# Is lifestyle coaching a potential cost-effective intervention to address the backlog for mental health counselling? A Rapid Review

**DOI:** 10.1101/2023.01.20.23284835

**Authors:** Abraham Makanjuola, Rachel Granger, Kalpa Pisavadia, Rhiannon Tudor Edwards

## Abstract

**Aim:** The aim of this rapid review was to investigate whether lifestyle coaching could provide a cost-effective alternative to counselling for the UK National Health Service (NHS) treatment of common mental health conditions such as stress, anxiety and depression.

**Methods:** A rapid review approach was used to determine the evidence of health economics evaluations in the field of mental health services. A literature search of PubMed, CINAHL, Cochrane Library, ASSIA, PsycINFO and MEDLINE produced 2807 articles. We removed 778 duplicates, and 2029 study articles remained. Two reviewers screened titles and abstracts (RG and KP), and 37 papers met the inclusion criteria of this review. Following a full-text screening, a further 27 papers were excluded due to lack of relevance. Study designs which did not include economic evaluations (n=15) or did not include an evaluation treatment of mental health conditions with talking therapies (n=15) did not meet the inclusion criteria. Ten papers were included in the final rapid review.

**Results:** The database search yielded study articles which focused on the cost-effectiveness of counselling and other talking therapies such as Cognitive Behavioural Therapy (CBT). No literature was found to determine the cost-effectiveness, or effectiveness of lifestyle coaching. Due to a lack of economic evaluations, this review could not determine the potential cost-effectiveness of lifestyle coaching as a means of addressing the backlog for mental health support such as counselling in the NHS.

**Conclusion:** This review highlights the research gap in assessing the cost-effectiveness of lifestyle coaching for treating common mental health disorders. The proposed next step is to evaluate the effectiveness and cost-effectiveness of lifestyle coaching versus current treatment as usual (counselling) by using a feasibility randomised control trial.

**Paper type:** A rapid review

**Article summary:** *Strengths and limitations of this study:*

- This rapid review found a range of different economic evaluations of mental health interventions for common mental health issues.
- All of the study articles found were moderate to high quality, some of the included study articles met all of the checklist criteria.
- This rapid review found no evidence from a UK study setting. However, all study articles came from OECD countries that share similar legal structures and policies with comparable populations.
- Despite being mentioned in a number of studies, it is unclear what treatment as usual refers to, and is perhaps not as usual as the studies suggest.

## Introduction

The World Health Organisation defines mental health as “a state of well-being in which the individual realises their abilities, can cope with everyday stresses of life, can work productively and is able to contribute to his or her community” [2]. The concept of mental health has changed in recent years, moving away from a purely clinical or medicalised approach to more focus on supporting people with mental disorders to cope well with life by developing self-care and self-management strategies [3]. This is reflected in the diversification in mental health service provision, with many current services moving away from traditional clinically based open-ended counselling approaches, to techniques which are more fixed-term, problem focused and action-orientated, such as Cognitive Behavioural Therapy (CBT). In more recent years the use of a coaching approach has also been used for managing mental health [4–6].

The use of coaching techniques within a sports or corporate environment are well established [7,8]. In recent years coaching techniques have also been applied to health and wellbeing [9]. Coaching in a corporate environment uses a short- to medium-term collaborative working relationship between a coach and client to identify and optimise the client’s personal strengths, recourses, positive states and behaviours. It uses a combination of goal setting and positive psychological approaches to facilitate the clients’ personal growth, functioning and wellbeing [10]. The definition of health and wellness coaching, also called lifestyle coaching, is still broad. However, similar to coaching in a corporate environment, the approach is based upon positive psychology and behaviour change theory, is patient-and goal-focused and often utilises established behavioural techniques, such as CBT and mindfulness-based interventions [11,12]. Although there is overlap with the aims and techniques of coaching and counselling, the underlying approaches are different (see figure 1). Lifestyle coaching is more collaborative, forward-thinking and goal orientated, with less focus on the intrinsic processing of feelings or unconscious behaviour addressed with counselling [13,14]. Although lifestyle coaching is not specifically designed to be used with non-clinical populations, research suggests that 25-50% of the general public who received lifestyle coaching had clinical levels of mental health conditions. More specific working populations have higher rates of clinically significant mental health conditions than the general public [14]. In addition, a meta-analysis of positive psychology studies, which included individual, group and self-help coaching interventions, showed a significant positive impact on individuals’ subjective and psychological wellbeing [15].

**Figure 1:**
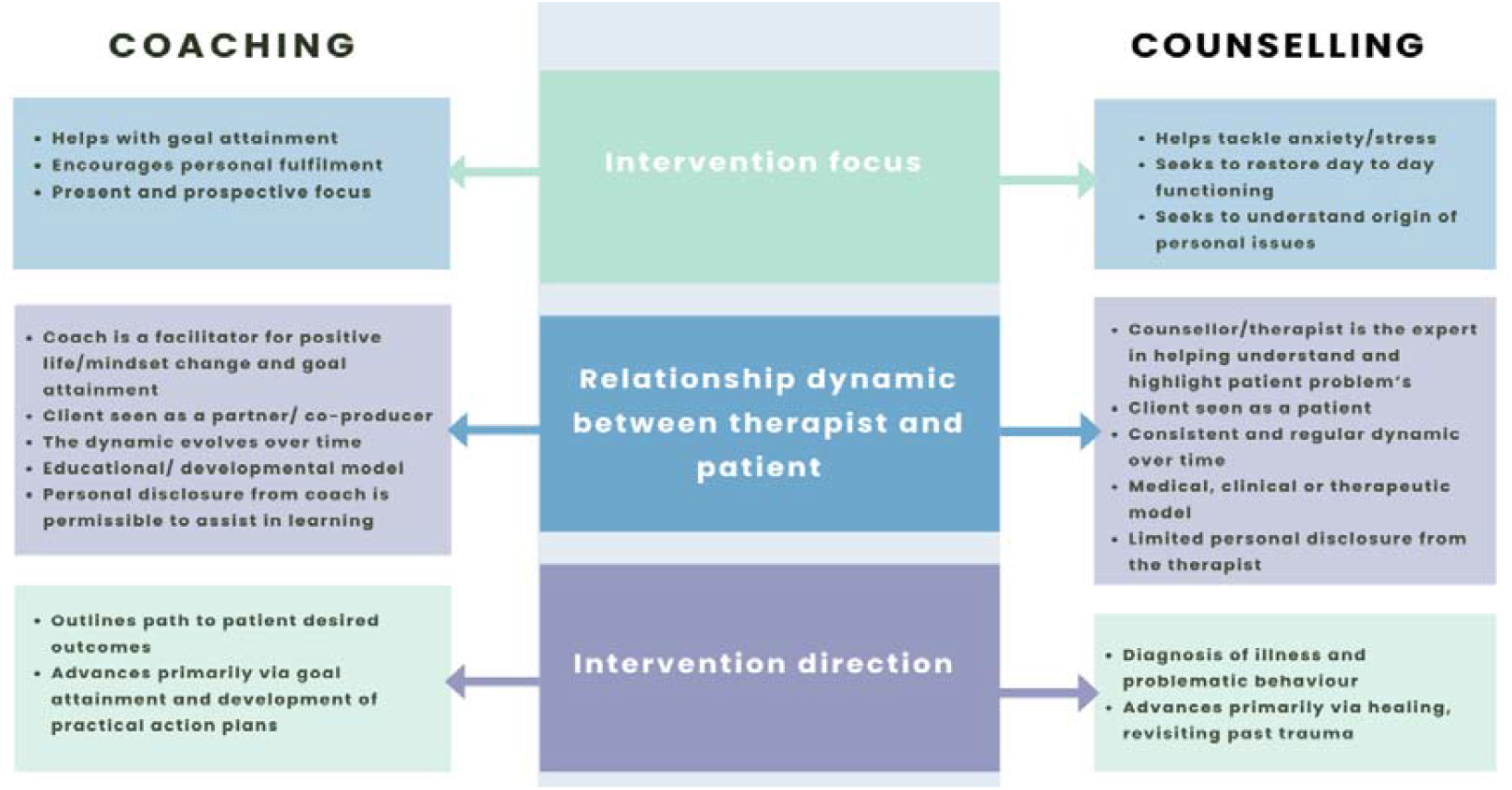
General differences between coaching and counselling [25].

There may also be other factors that would make the use of lifestyle coaching an effective treatment for mental health conditions. Lifestyle coaching is often shorter-term as it is focused on specific goals, compared to a more open-ended physiotherapy approach [14]. Shorter treatment periods may unlock staff resources and enable more people to be treated over the same time period. Lifestyle coaching does not have the stigma that can be associated with more traditional treatment for a mental health condition, so it may reduce barriers to seeking help [16]. The training process for lifestyle coaches is considerably shorter than the years of training to become a psychotherapy practitioner, which would make it quicker to recruit more lifestyle or recovery coaches.

With the inexorable rise of people living with long-term conditions, the National Health Service (NHS) is moving towards making self-care a priority, supporting people to manage their conditions, make informed treatment choices and avoid complications [17]. Although Health Coaching is one of the key NHS interventions to manage self-care, much of the evaluation work to date has focused on programme design, implementation and process evaluation, rather than evaluating the effectiveness and cost-effectiveness of treatment [18]. Therefore, the aim of this rapid review was to assess whether there was evidence for the cost-effectiveness of lifestyle coaching interventions for common mental health disorders that could support the NHS health coaching approach.

Common mental health disorders (CMD) include depression, generalised anxiety disorder, panic disorder, obsessive-compulsive disorder (OCD) and post-traumatic stress disorder (PTSD). Although there is considerable variation in the severity of CMD, many of these disorders are associated with significant long-term disability and often with ongoing cycles of relapse and remission. Prior to the COVID-19 pandemic around one in six adults (17%) met the criteria for a common mental health disorder [19,20].

The National Institute of Health and Care Excellence (NICE) CCG41 indicator guidance for people suffering from depression and anxiety states that talking therapies are important to recovery for all ages but particularly for those over 65 due to a lack of representation in services for this demographic [21]. NICE guidelines for CMD provide a number of treatment pathways patients can choose from in collaboration with their healthcare professional. An independent NICE guideline committee developed a guideline to treat and manage depression in adults. Patients suffering from less severe depression can include both the use of psychotropic medication as well as psychological treatments, including CBT, interpersonal therapy (IPT), behavioural therapy, psychotherapy, self-help and support groups in their initial treatment [22]. Despite psychological interventions generally being preferred by patients, due to limited availability, the most common treatment method for CMD in primary care is medication [23].

Although most of the evidence used to develop the NICE guidelines was mental health-specific (focusing on depression), general NICE guidance for improved treatment of common mental health disorders focuses on improved access to services, including local care pathways in a range of settings and alternative modes of delivery (for example, telephone and online services). The single economic study included in the NICE evidence review reported that a more formalised stepped care CBT treatment for adults with common mental health disorders resulted in a higher number of Quality Adjusted Life Years (QALYs) at a lower cost than the more open-ended care as usual [24].

### Aim

To review the evidence on whether lifestyle coaching is a cost-effective alternative to counselling for mental health within the NHS and the UK.

### Objectives

1. Identify current lifestyle coaching interventions for mental health
2. Identify the differences between lifestyle coaching and counselling
3. Determine whether lifestyle coaching is a cost-effective alternative to counselling for mental wellbeing

### Rapid Review Methodology

This study adopted a rapid review approach abridging some sections of the standard systematic review process to generate quality evidence in less time. This methodology follows the minimum requirements for rapid reviews, featuring a study protocol, a systematic database search, study screening, data extraction, critical appraisal, and narrative synthesis [26]. This revised methodology is used by the Wales COVID-19 Evidence Centre (WCEC) [27–29]. The review protocol was not registered on a review protocol database.

### Patient and Public Involvement

No patient involvement.

### Search Strategy

The key evidence sources of this rapid review include PubMed, Cumulative Index to Nursing and Allied Health Literature (CINAHL), Cochrane Library, Applied Social Sciences Index and Abstracts (ASSIA), PsycINFO and MEDLINE. The searches were conducted on 23^rd^ January 2022 using a five-year timeframe from January 2017 to January 2022. Search terms available in (See Appendix 1).

The Mendeley Desktop reference management software was used to manage study articles found and remove duplicates.

The inclusion criteria was based on the PICO approach (shown in Table 1) and consisted of peer-reviewed economic evaluations of coaching, mentoring and counselling therapies from Organisation for Economic Co-operation and Development (OECD) countries in English and French (AM is able to read French texts) published from 22^nd^ January 2017-21^st^ January 2022 [30]. Therefore, the exclusion criteria consisted of non-peer-reviewed economic evaluations of coaching, mentoring and counselling therapies from non-OECD countries which were also not written in English or French. Those papers that were not published within the five-year timeframe were also excluded.

**Table 1:** PICO Framework

| Review question |  |
| --- | --- |
| Is lifestyle coaching a cost-effective intervention to address the backlog for mental health counselling? |  |
| Participants | Economic evaluations for lifestyle coaching and counselling<br>Cost-Benefit Analysis<br>Cost-Utility Analysis<br>Cost-effective Analysis<br>Cost effective*<br>Social Return on Investment<br>Return on Investment<br>Social Cost-Benefit Analysis<br>Cost-Minimization Analysis<br>Cost-Consequence Analysis<br>Cost-effective alternative to counselling for mental health in the NHS |
| **Intervention / exposure** | Lifestyle coaching and similar talking therapies.<br>Coaching<br>Lifestyle coaching<br>Mentoring<br>Therapy<br>Talking Therapy |
| **Comparison** | Counselling |
| **Outcomes** | Mental Wellbeing<br>Mental Health<br>Confidence<br>Wellbeing<br>Relationship*<br>Goal attainment<br>Self-management<br>Self-efficacy<br>Social Value |
|  | ICER (Incremental Cost-effectiveness Ratio)<br>ICUR (Incremental Cost-Utility Ratio)<br>Unit*<br>QALY (Quality Adjusted Life Year)<br>DALY (Disability Adjusted Life Year)<br>HRQoL (Health Related Quality of Life)<br>QoL (Quality of Life) |
| **Other Study Considerations** |  |
| Primary, secondary, grey literature, preprints |  |

### Selection of Studies

Two reviewers (RG and KP) independently selected potentially eligible studies in the first phase, where selection was based on titles and abstracts. In the second stage the full texts of selected studies were accessed for eligibility. All studies identified as potentially eligible by at least one of the reviewers were re-assessed by RG and KP, in conjunction with a third reviewer (AM) who produced a consensus on the final list for phase two.

### Data Extraction

Data extraction and study quality assessment were performed by all three reviewers (AM, RG, KP). Data was collected on country, study design, intervention type, data collection methods and dates, sample size, type of participants, and primary and secondary (see Appendix 2).

### Quality Assessment

Quality assessment was carried out by all members of the review team (AM, RG, and KP) using the Joanna Briggs Institute (JBI) critical appraisal tools: Economic Evaluation, Systematic Review and research syntheses checklist, and Randomised clinical trial checklist (see Appendix 3).

## Results

The database search yielded 2807 study articles, 778 of which were duplicates. The remaining 2029 titles and abstracts were screened, with 37 study articles meeting the criteria for this review. Following a full-text screening, a further 27 papers were excluded due to lack of relevance. Study designs that did not include an economic analysis (n=12) or studies that did not include interventions to treat common mental health conditions such as talking therapy (n=15) did not meet the inclusion criteria. Following full text screening of the remaining results, ten study articles were included in the final rapid review (See Figure 2).

**Figure 2.**
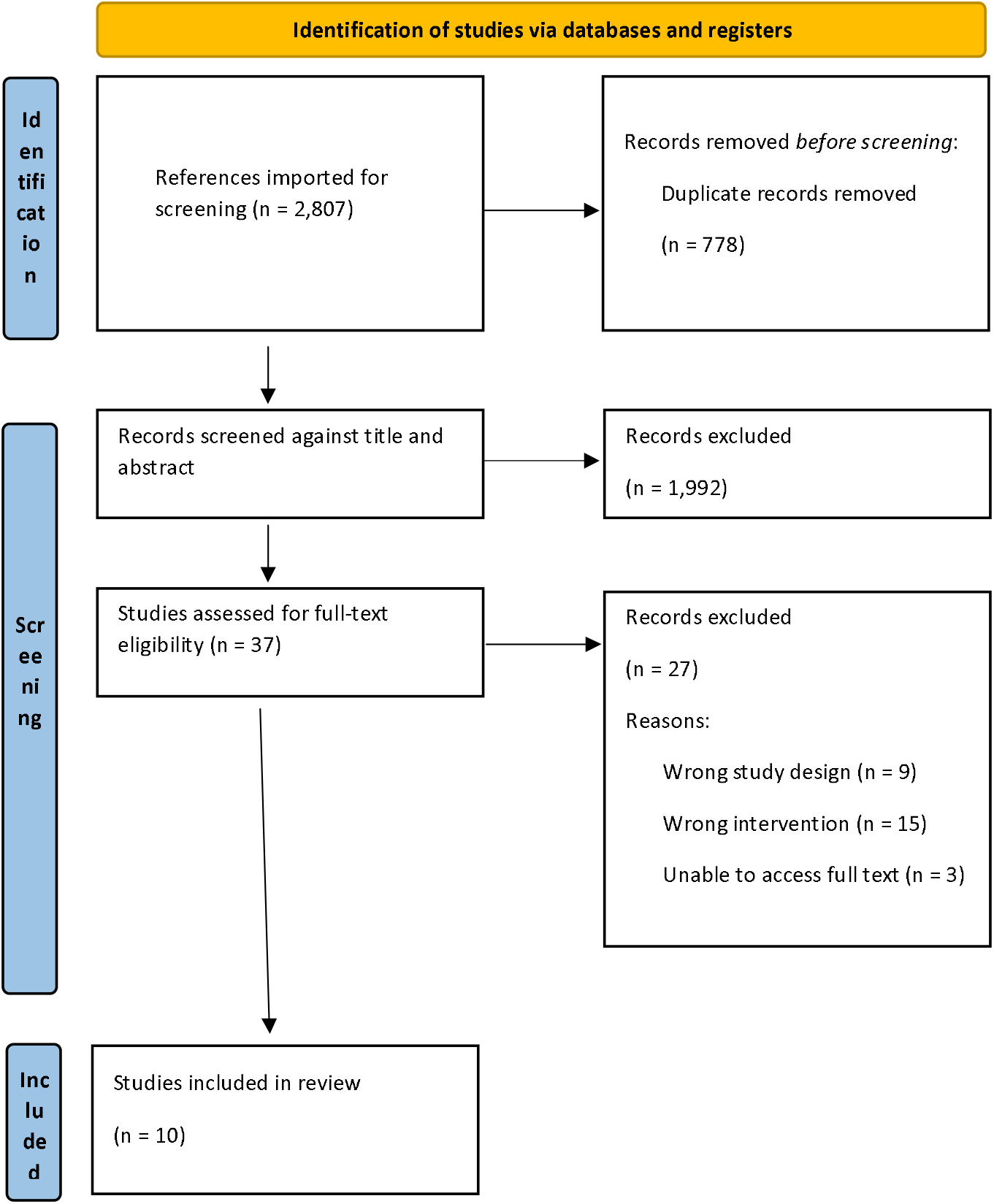
PRISMA study selection flowchart (Page *et al*>, 2021)

**Figure 3:**
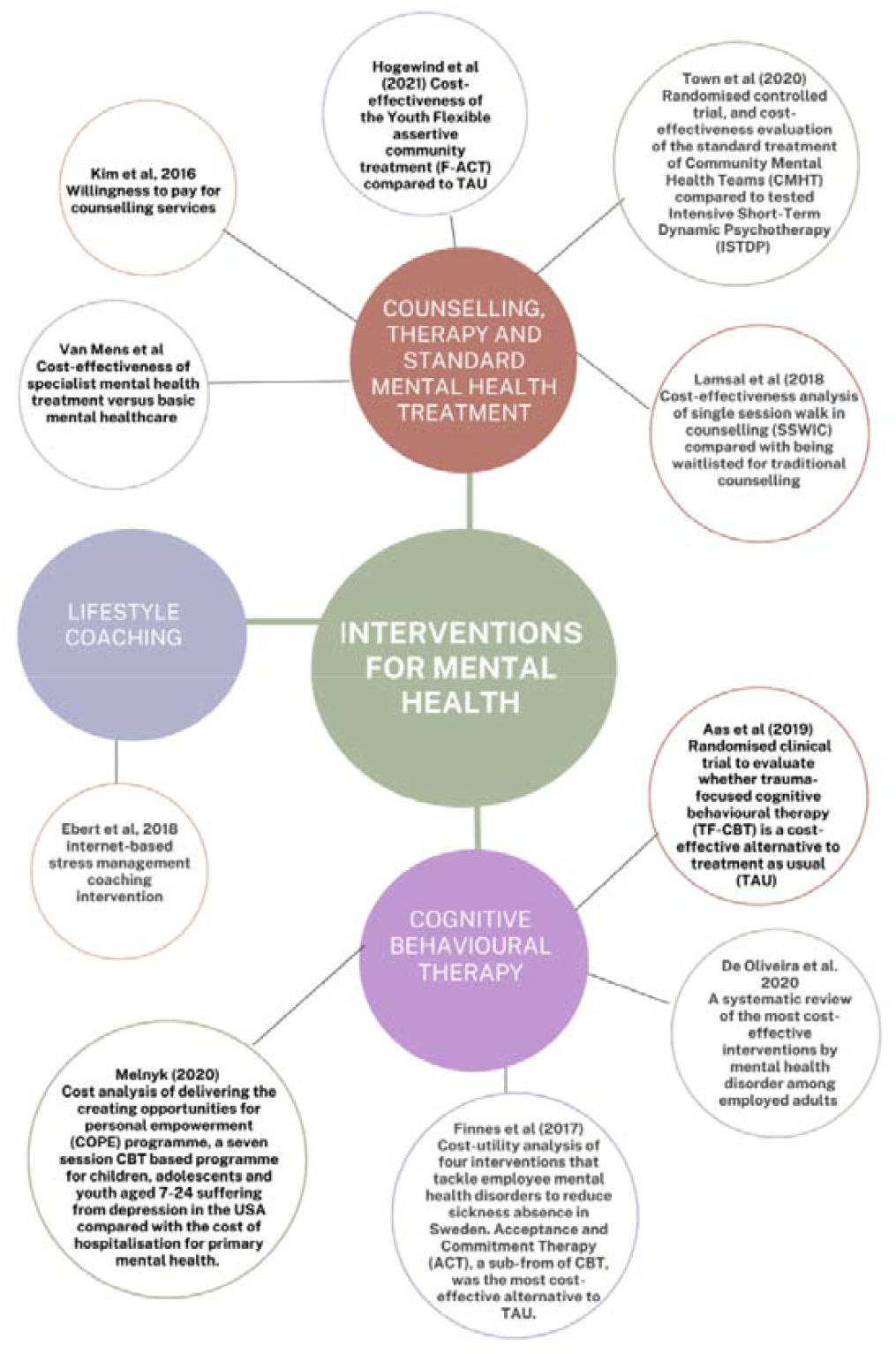
Evidence divided by study intervention type.

All ten study articles included in this rapid review were peer-reviewed. Nine of the study articles were primary studies, and the methodologies of the included studies were randomised controlled trial (RCT) (n=4), economic evaluation (n=3), quasi-experimental design (n=2), and a systematic review (n=1).

The Economic evaluations included in this rapid review consist of Cost-effectiveness analysis (n=7), cost-benefit analysis (n=1), cost-utility analysis (n=1) and stated preference using the contingent valuation method (n=1).

All ten included study articles are of a moderate to high quality, according to the JBI quality appraisal checklists [31–40] (See Appendix 3).

This rapid review found no evidence of economic evaluation of coaching or counselling interventions in Wales or the rest of the UK. The countries included in this rapid review were from the OECD. Countries included Netherlands (n=3), USA (n=2), Canada (n=2), Norway (n=1), Germany (n=1), Sweden (n=1), and South Korea (n=1).

One study article relates to the theme of coaching for mental health disorders [33].

Four study articles relate to the theme of CBT [31,34,38,41], and five study articles relate to the theme of Counselling, therapy and standard mental health treatment [35–37,39,40].

Five of the study articles are in a workplace setting [33,34,36,37,41]. Although this review did not find economic evidence for the cost-effectiveness of lifestyle coaching, this review demonstrates that similar approaches using talking therapies such as CBT are cost-effective treatments for some mental health disorders. The following discussion on the papers included give a more detailed overview of findings.

### Coaching

Ebert et al (2018) conducted a cost-effective and cost-benefit analysis alongside a randomised controlled trial of an internet-based stress management coaching intervention for employees of a health insurance company in Germany [33]. The findings indicate that the (guided) internet and mobile-supported occupational stress-management intervention (iSMI) was cost-effective and led to cost savings for employers within the first six months post-randomisation compared with the treatment as usual control group. The CBA yielded a net benefit of €181 [95% confidence interval (CI)] per participant.

### Cognitive Behavioural Therapy (CBT)

Aas et al (2019) conducted a randomised clinical trial to evaluate whether trauma-focused cognitive behavioural therapy (TF-CBT) is a cost-effective alternative to treatment as usual (TAU) for participants in Norway [31]. Aas et al found that TF-CBT is likely to be a cost-effective alternative to TAU for youth living with PTSD and that patient usage of other services such as welfare services, medication and school nurses do decline.

A systematic review conducted by de Oliveira et al. to provide a knowledge base of the most cost-effective interventions by disorder among employed adults with mental health disorders found that CBT is cost-saving and, in some cases, cost-effective in tackling depression [41]. Further results indicate that addressing workplace mental health will improve wellbeing and productivity and could improve profits.

Finnes et al (2017) conducted a cost-utility analysis of four interventions that tackle employee mental health disorders to reduce sickness absence in Sweden [42]. Acceptance and Commitment Therapy (ACT), a sub-form of CBT, was the most cost-effective alternative to TAU.

Melnyk (2020) conducted a cost analysis of delivering and creating opportunities for personal empowerment (COPE) programme [38]. COPE is a seven-session CBT based programme for children, adolescents and youth aged 7-24 suffering from depression in the USA compared with the cost of hospitalisation for primary mental health. Melnyk found that COPE is a cost-saving intervention for teens, adolescents and youth living with depression. $14,262 (USD) is saved for every hospitalisation that is averted.

#### Counselling, therapy and standard mental health treatment

Hogewind et al (2021) evaluated the cost-effectiveness of the Youth Flexible assertive community treatment (F-ACT) compared to TAU in the Netherlands [35]. The results of the retrospective cost questionnaire indicate that F-ACT resulted in higher quality of life among adolescents. The Incremental Cost-effectiveness ratio shows that F-ACT is more effective than TAU for youth, but it is associated with higher costs.

Kim et al (2016) conducted an economic evaluation using the Contingent Valuation Method (double-bounded dichotomous choice questions) to elicit the willingness to pay (WTP) of South Korean university students and office workers for counselling services [36]. Kim et al found that those earning a monthly salary of KRW 2,500,000 ($2223.21 USD in 2016) and equally paid KRW 66,625 ($59.25 USD in 2016) per month for medical insurance had a WTP of a 1% premium to access more counselling. Prior experience of counselling and disposable income positively affected participant WTP.

Lamsal et al (2018) conducted a cost-effectiveness analysis of single session walk in counselling (SSWIC) compared with being waitlisted for traditional counselling. The clients (16 years and older) of two community-based family services in Canada [37]. SSWIC is not cost–effective when compared to traditional counselling. However, SSWIC provides a service that can alleviate pressure on the Canadian mental health system by reducing waitlist times for long term counselling services.

Town et al (2020) conducted a randomised controlled trial, and cost-effectiveness evaluation of the standard treatment of Community Mental Health Teams (CMHT) compared to tested Intensive Short-Term Dynamic Psychotherapy (ISTDP) of adult patients with treatment-resistant depression in Canada [40]. Standard treatment consists of counselling, CBT and group therapy. Town et al reported minimal difference in general anxiety and wellbeing scores between the two groups; however, the ISTDP group displayed lower cost and higher quality of life. When the data of high-volume patients were removed, the differences in cost were minimal.

Van Mens et al conducted a quasi-experiment to evaluate the cost-effectiveness of specialist mental health treatment versus basic mental healthcare of patients experiencing common mental health issues who had not previously received mental health treatment in the Netherlands. Patients were compared from pre and post introduction of basic mental healthcare [39]. The average unit cost for specialised mental healthcare is €75 and, for basic mental healthcare it is €83. The average number of treatment hours is 39.4 in specialised mental healthcare and 9.8 in basic mental healthcare. Results indicate that basic mental healthcare is cost-saving; the group receiving basic health care experienced similar health outcomes as the group receiving specialist treatment, for on average €2132 (plll<lll0.001) less.

## Discussion

This rapid review set out to determine whether lifestyle coaching is a cost-effective alternative to counselling for mental health conditions within the NHS, particularly in light of the backlog or of referrals post-pandemic. Despite a comprehensive search strategy, limited evidence of economic evaluations of lifestyle coaching for mental health were found. This rapid review also found no economic evaluations of mental health interventions in the UK. However, the included economic evaluations were from OECD countries which are comparable to the UK [43].

Treatment/Therapy as usual (TAU) was mentioned frequently as a comparator in the cost analyses. However, TAU intervention was either inconsistent or not mentioned throughout the different study settings. Examples of TAU include:

○ 53 minutes sessions using therapy strategies which are client-centred, psychodynamic, family and cognitive intervention [31].
○ Any intervention offered by a primary care centre or care facility [42].
○ Care from a multidisciplinary team comprised of a psychologist, psychiatrist, family therapist and social workers [35].

Positive recovery-focused therapies, including recovery coaching (a type of lifestyle coaching focused on mental health) have also begun to be used to treat mental health disorders in recent years [44–46]. This approach has been already used in one NHS trust in Devon [47]. The use of CBT, mindfulness-based therapies, and self-help interventions (all of which can be used in lifestyle coaching) have already been identified as effective treatments for common mental health disorders [23]. In addition, a meta-analysis of randomised control trials reported that specific positive psychology interventions, including lifestyle coaching, solution coaching and self-management techniques, can enhance subjective and psychological well-being and reduce depressive symptoms [15]. It must be acknowledged that there are limitations to using a coaching approach for mental health. The effectiveness of coaching is dependent on people’s motivation and desire to improve areas of their life [47]. People with severe mental health problems may feel stuck in their current state, not feeling able to act positively within their problem situation [48]. Shechtman et al (2019) conducted a 12-week intervention comparing two groups of Israeli parents of children with attention deficit hyperactivity disorder (ADHD). One group attended weekly coaching sessions with a trained coach, while the other read a self-help book with coaching techniques [49]. Both parental groups reported higher competence and efficacy. However, the group that attended therapy sessions reported a greater improvement in coping with children’s negative emotions than the control group. The drop-out rate for the self-help intervention was over twice that of the coaching intervention (23% and 10%, respectively). This discrepancy in drop-out rates suggests that one-to-one coaching is more effective than self-guided study. Participants are more likely to adhere to the full course with one-to-one coaching.

Six of the ten study articles included in this rapid review evaluated the cost-effectiveness of CBT for common mental health conditions. Traditional psychotherapy and counselling are based on the medical/clinical model of practice that relies on diagnosis and pathology [50]. This approach regards the person as a “patient” and focuses predominantly on resolving problems that have already been established. Although this methodology is still used with severe mental health conditions, many counselling approaches have moved away from the “patient” approach in recent years. For example, the technique of Cognitive-Behavioural Therapy (CBT) focuses on specific symptoms or problems and tends to be more time-limited than traditional approaches [51]. There is also the sub-discipline of Coaching Psychology which utilises established psychological theories and models of lifestyle coaching to enhance well-being and performance [52].

There have been concerns raised about the lack of professional-wide training or clear governing practice of coaching for mental health. Although a valid concern, a recent scoping review to address these concerns reported positive outcomes for the use of coaching for mental health difficulties [4].

Common mental health disorders, such as depression, stress and anxiety, are associated with a range of symptoms that often lead to adverse health, social and economic consequences for the individual affected and society [53]. Prior to the pandemic, the lack of service provision for specialised mental healthcare within the NHS and the need to develop alternative approaches had already been highlighted [54]. The initial and long-term effect of the pandemic on people’s wellbeing and mental health has been widely reported throughout the UK. The level of mental health issues almost tripled in the first few months of the pandemic (rising from 12% pre-pandemic to 28%), with the numbers of problems reported and the proportion of severe problems still significantly higher than pre-pandemic levels [55]. To deal with this increased demand, the need to implement effective alternative strategies to manage demand for support for mental health conditions mental healthcare within the NHS is now imperative.

### Workplace interventions

There is growing evidence that there are economic gains to be made from workplaces addressing mental disorders amongst employees. Absenteeism and presenteeism are significant cost drivers that often exceed the cost of treatment [33]. Common mental health disorders such as depression, anxiety and severe stress are the leading causes of sickness absence in most high-income countries. The total cost of sickness absence to society has been estimated to be 3% to 4% of the gross domestic product in European countries [34]. CBT has been shown to be an effective early intervention for depression, reducing absenteeism and presenteeism and thus, providing a positive return on investment. A systematic review conducted by de Oliveira et al, found that every dollar invested in workplace CBT interventions with care management produced a return on investment varying between $0.39 and $3.35 (USD in 2020) per employee after one year [56]. The global audit company Deloitte found that increased technology usage has had detrimental effects on mental health [57,58]. Poor mental health in employee costs UK employers £29 billion a year in reduced productivity as a result of presenteeism. Presenteeism describes a working culture whereby staff are present at work but are not working at their best [59]. The COVID-19 pandemic has led to more digital presenteeism whereby staff feel obliged to always be reachable via digital platforms such as Teams, Outlook and Slack [58]. Deloitte found through their report that for every £1 invested in employee mental health there is an average return of £5 [57]. There is evidence that CBT is more cost-effective than pharmacotherapy [60]. However, despite psychological interventions generally being preferred by patients, due to limited availability, the most common method of treatment for CMD in primary care is medication [23].

### Remote Interventions

One of the greatest changes in mental health treatment has been the move to online treatment during the pandemic, both for traditional open-ended psychological interventions [61] and shorter-term programmes specifically utilised for the pandemic [62]. A previous meta-analysis has shown that remote interventions can be effective in improving psychological wellbeing [15]. There is also evidence that remote computer-delivered CBT programmes (with reduced therapist contact) are as clinically effective and more cost-effective than standard CBT treatments [63]. However, a recent review reported that prior to the pandemic, there was little evidence for large-scale implementation of or the long-term impact of remote services [64]. This paper also raised the issue of digital exclusion for some sectors of society. Although online services are likely to be more cost-effective, the changes enforced by the pandemic give us ample opportunity to review both the online effectiveness and cost-effectiveness of remote mental health services and the impact on different sectors of society.

### Current NHS Approaches and Waiting Times

Prior to the pandemic, the approach of NHS mental health services was already changing to address service availability and access issues. A decade ago, NHS England introduced Improving Access to Psychological Therapies (IAPT) to improve access to psychotherapy for the general population. The programme, which follows NICE recommendations, is designed to treat CMD, including anxiety and depression [65]. IAPT also uses digitally enabled therapy, enabling therapy content to be delivered online or through mobile applications. The approach is focused on self-study, with additional support and reinforcement by trained therapists. Although this move to using more accessible services has provided additional access and support for many patients, recent waiting time figures show that the current mental health service provision, including IAPT, is still not sufficient to meet post-pandemic needs [66]. The ability to access services within six weeks has shown a continuous downward trend (with a further 3% decrease over the 12 months of Feb. 2021-2022), although access to services within 18 weeks has remained relatively stable. With a predicted increase in demand for mental services, there is the potential to investigate whether the NHS could provide additional interventions including lifestyle coaching or a similar service in the workplace for mental health disorders [1].

#### Implications and Proposals for future research

Although several studies compare interventions such as CBT, Acceptance and Commitment Therapy (ACT) and workplace dialogue intervention (WDI), with treatment as usual such as counselling and psychotherapy, no studies compare lifestyle coaching as an alternative. The exclusion of lifestyle coaching within economic evaluations for mental health may mean that only a partial assessment of cost-effectiveness is undertaken, leading to sub-optimal resource allocation decisions. Research directed in this area in the UK will assist policymakers in finding alternatives to deal with the backlog that affronts the NHS. Additionally, future research should seek ways to expand the evaluative space of economic evaluations and explore approaches to integrate lifestyle coaching within various settings, such as schools, workplaces, and academic institutions.

This paper acknowledges the research gap for evidence for the effectiveness and cost-effectiveness of a lifestyle coaching intervention to treat CMD. Therefore, we propose a feasibility 2-pathway randomised control study (RCT) to directly compare current NHS service provision such as IAPT with an established coaching intervention.

## Conclusion

This rapid review found that CBT is cost-effective in dealing with common mental health issues such as PTSD, depression, anxiety and stress among a number of different populations and study settings. Incorporating CBT as an intervention for mental health conditions has the potential to reduce the need for medication, welfare and support services. The overlapping techniques of coaching for common mental health issues is more goal focused and aligns with the NHS’ Universal Personalised Care framework which fosters a co-productive intervention for treating common mental health issues.

This rapid review also demonstrates that online coaching as a stress-management intervention within occupational healthcare is cost-effective and cost-saving. However, there is little evidence on whether lifestyle coaching is a cost-effective alternative to available treatments with the potential to alleviate the pressures NHS mental health currently faces. To address this, we propose a feasibility RCT study approach that would enable the evaluation of both the effectiveness and cost-effectiveness of lifestyle coaching to address the current mental health care services backlog.

## Data Availability

All data produced in the present work are contained in the manuscript

## Acknowledgements

The authors thank the Bangor Institute of Medical and Health Research (BIMHR) team for their input into the structure and development of this review. We would also like to thank Yasmin Noorani, Academic Support Librarian at Bangor University, for her assistance in creating our search strategy. Additional thanks to Dr Catherine Lawrence for input and feedback on this paper.

## Contributors

The review was conceived by AM, RG, KP and RTE, and the protocol was developed by AM, RG and KP; searches were undertaken by AM and RG; article screening was carried out by RG and KP with mediation by AM; quality appraisal was undertaken by AM, RG and KP; data were interpreted by all authors; the manuscript was drafted by AM and RG and critically reviewed by all authors.

## Ethical approval

This study does not involve human participants.

### Appendix

#### Appendix 1: Search Strategy and terms

(Coaching[Title/Abstract] OR “Lifestyle coaching”[Title/Abstract] OR “life coaching”[Title/Abstract] OR “health coaching”[Title/Abstract] OR Mentoring[Title/Abstract] OR Mentors[Title/Abstract] OR Mentor[Title/Abstract] OR “Talking Therapy”[Title/Abstract] OR “talking therapies”[Title/Abstract] OR counselling[Title/Abstract] OR counseling[Title/Abstract] AND ((y_5[Filter]) AND (english[Filter] OR french[Filter]))) AND ((economic evaluation OR economic analysis OR cost analysis OR “cost-effectiveness” OR “cost-benefit” OR “cost-utility” OR “social return on investment” OR “social cost-benefit analysis” OR “Cost-Minimization Analysis” OR “Cost-Consequence Analysis” AND ((y_5[Filter]) AND (english[Filter] OR french[Filter]))) AND (Mental Wellbeing[Title/Abstract] OR “Mental Health”[Title/Abstract] OR Confidence[Title/Abstract] OR Wellbeing[Title/Abstract] OR Well-being[Title/Abstract] OR “Well being”[Title/Abstract] OR Relationships[Title/Abstract] OR “Goal attainment”[Title/Abstract] OR Self-management[Title/Abstract] OR Self-efficacy[Title/Abstract] OR “Social Value”[Title/Abstract] OR ICER[Title/Abstract] OR ICUR[Title/Abstract] OR Unit[Title/Abstract] OR QALY[Title/Abstract] OR DALY[Title/Abstract] OR HRQol[Title/Abstract] OR Qol[Title/Abstract]) AND ((y_5[Filter]) AND (english[Filter] OR french[Filter]))) Filters: in the last 5 years, English, French

### Appendix 2: Data extraction table

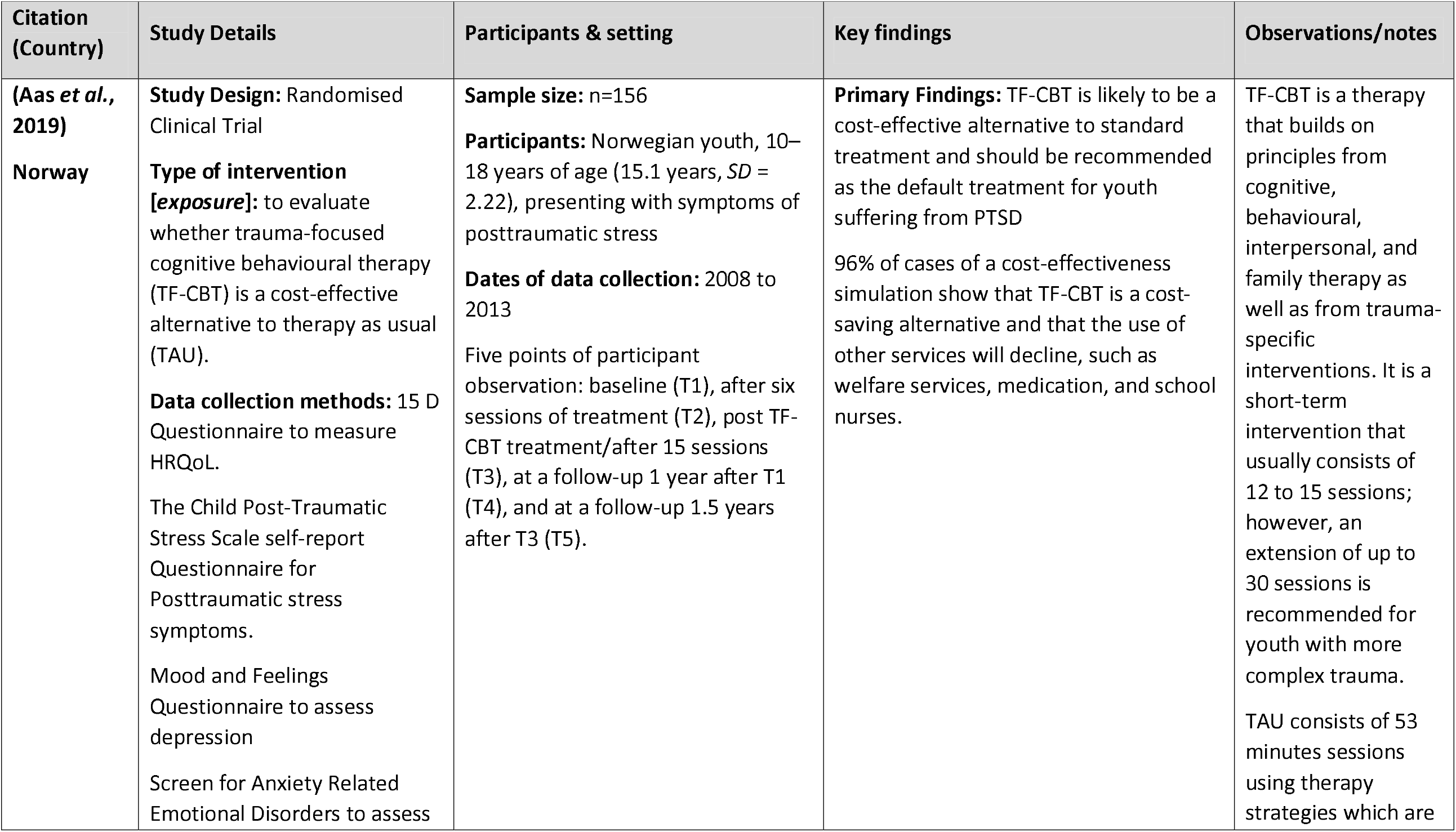
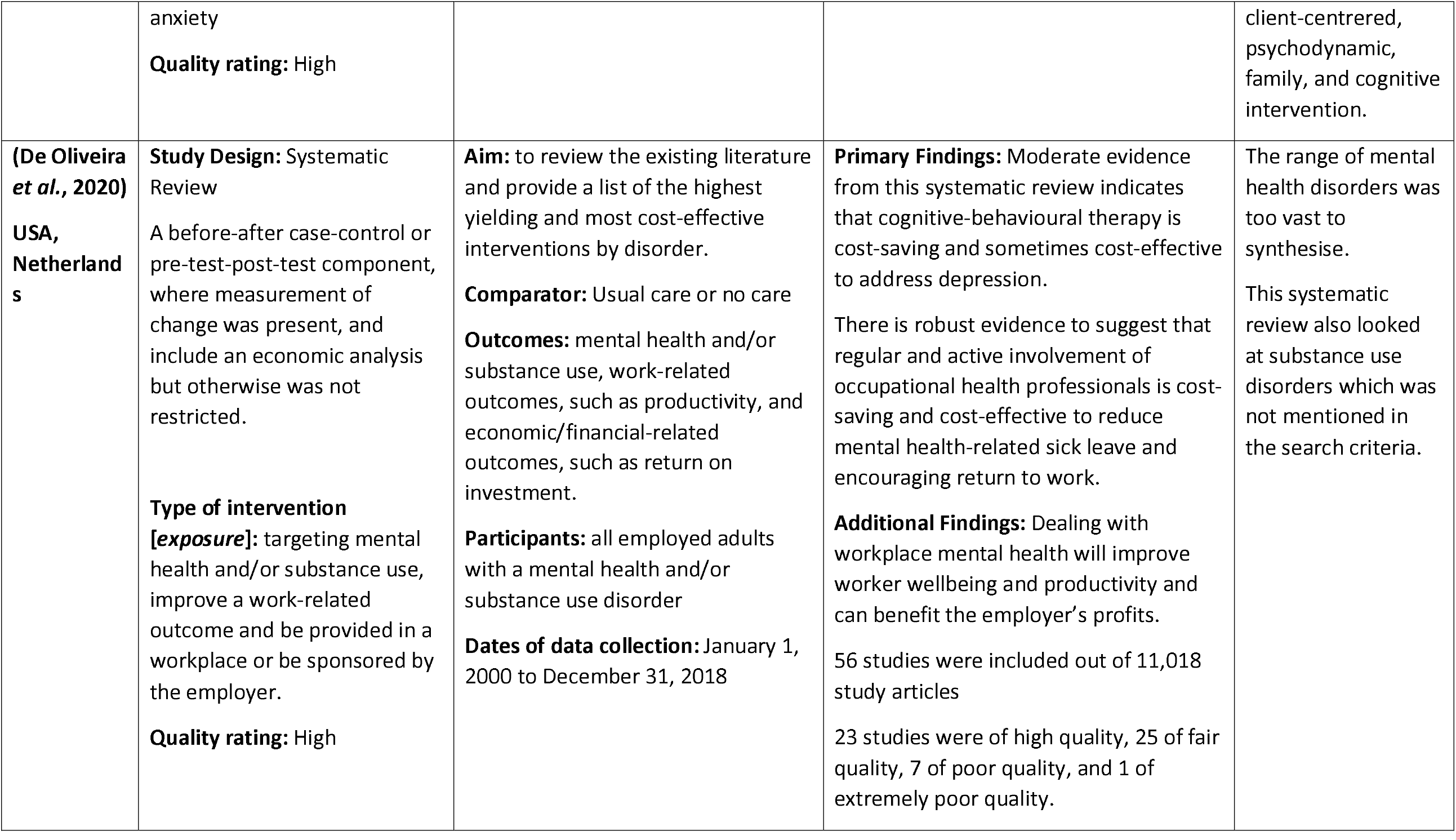
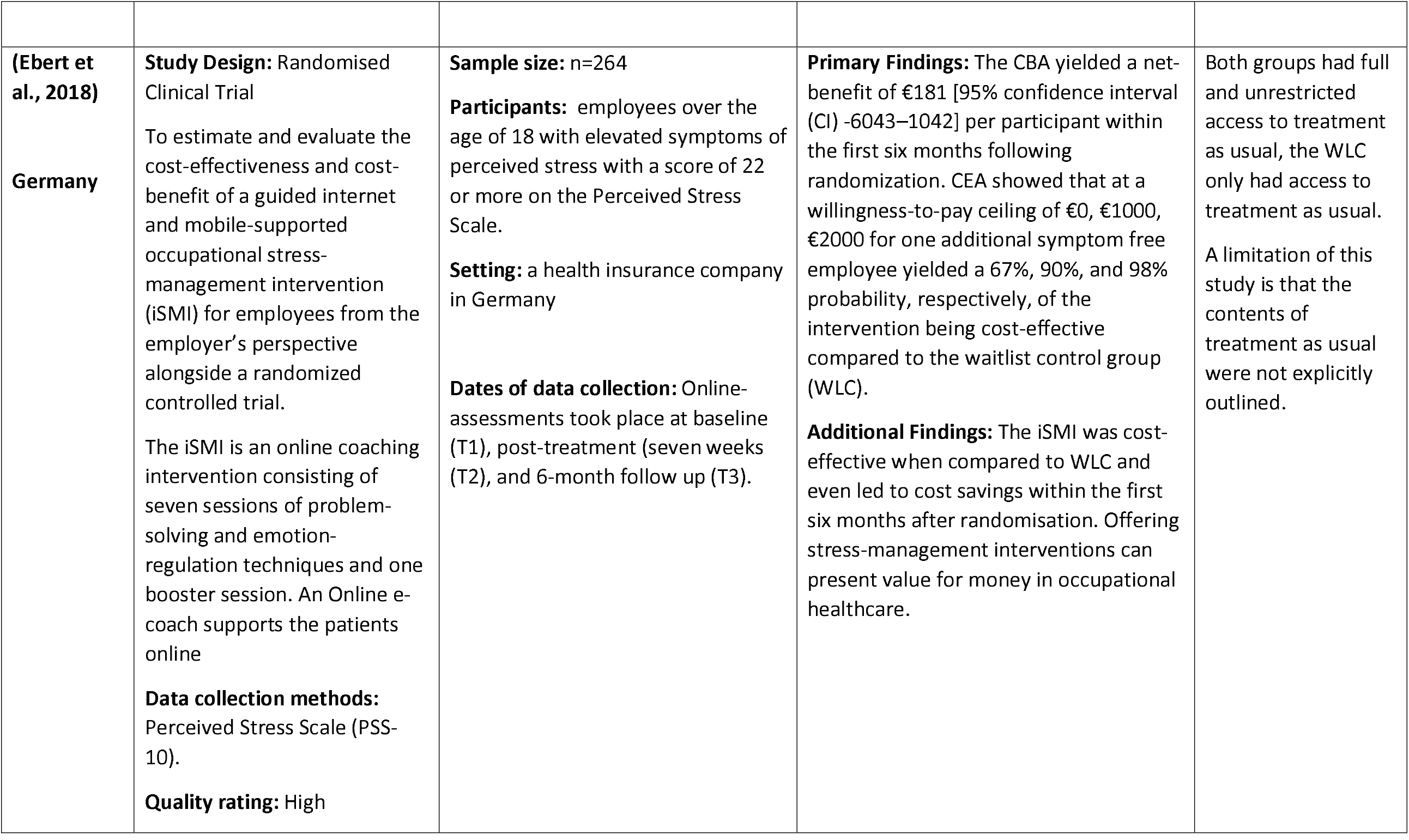
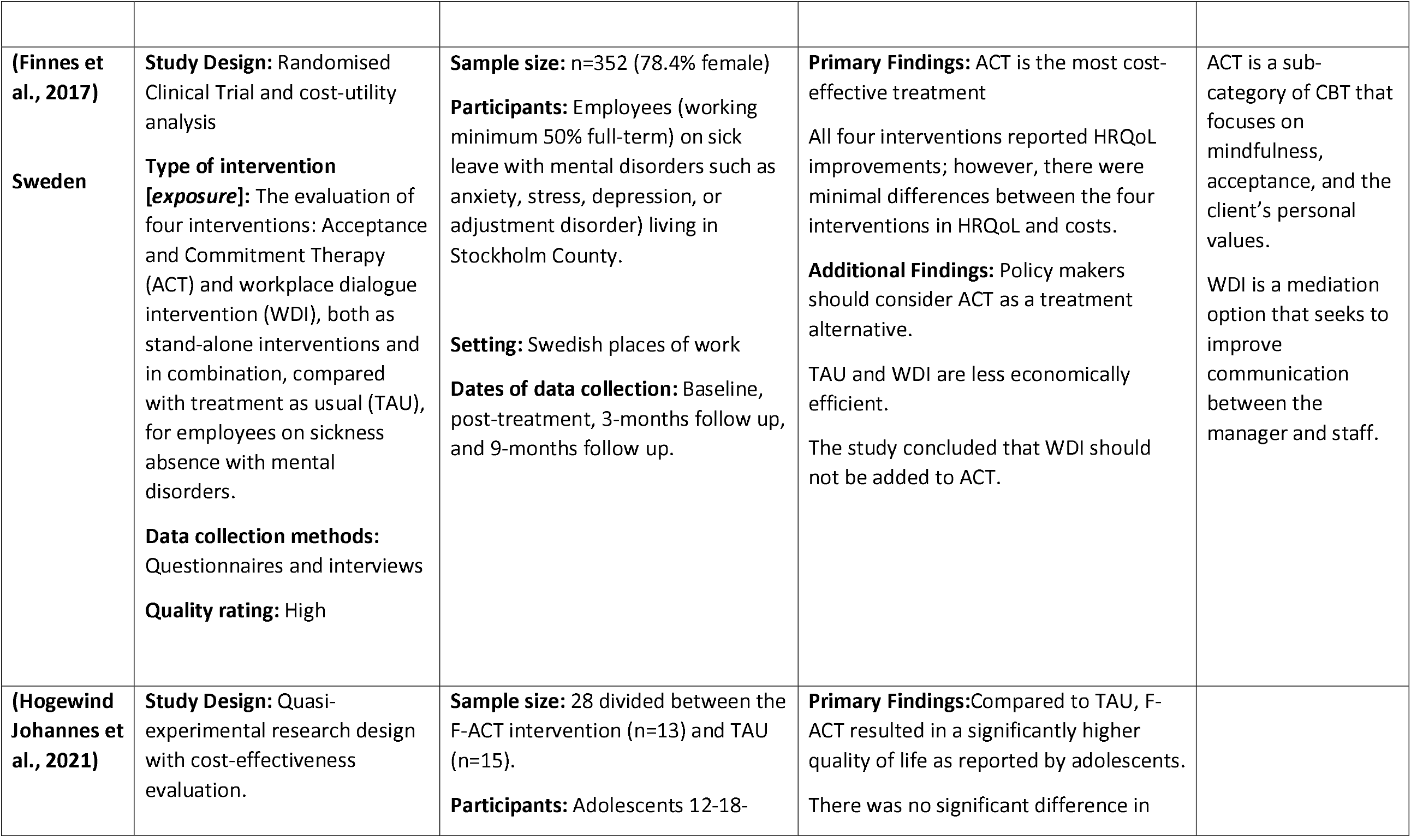
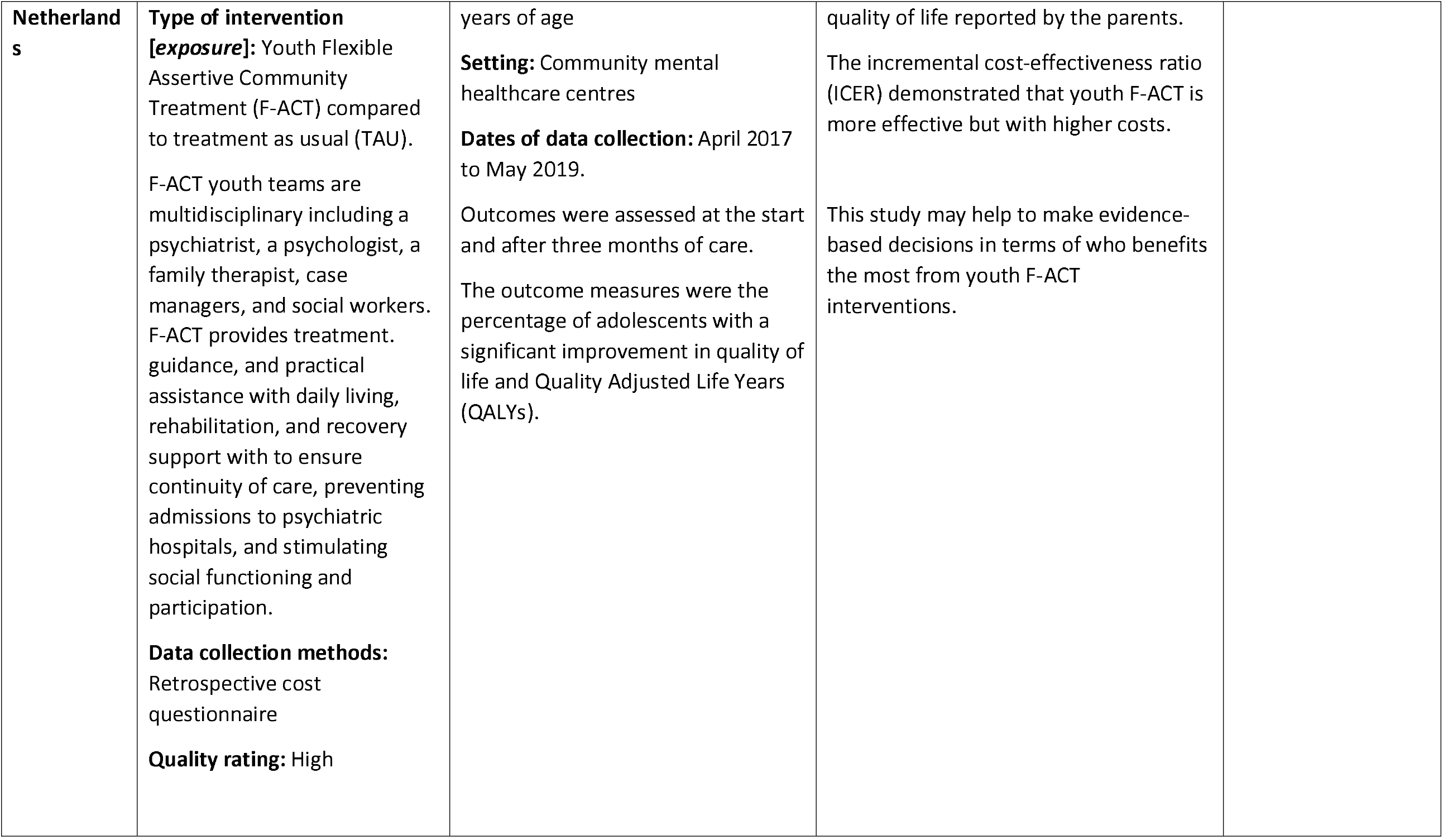
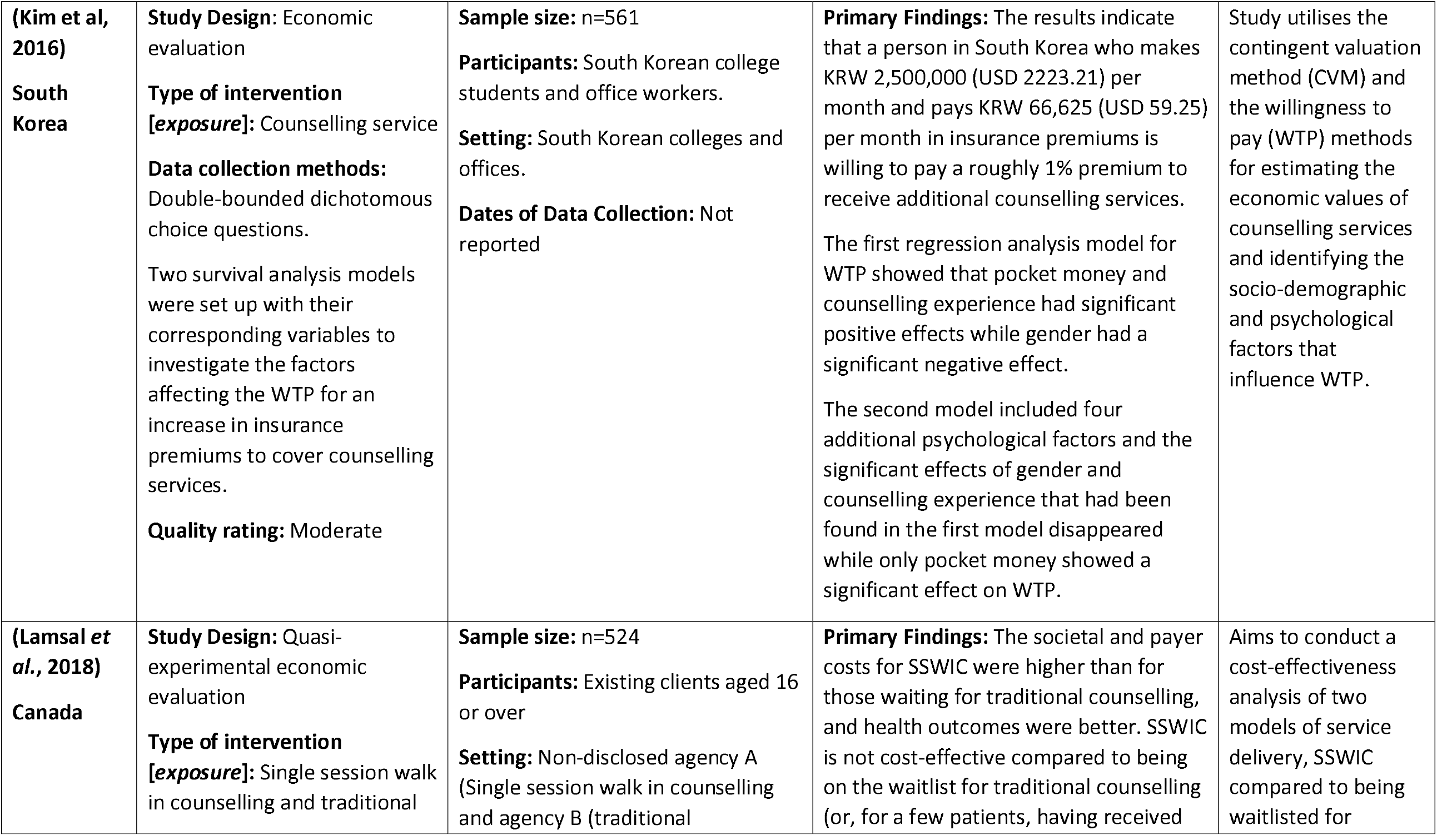
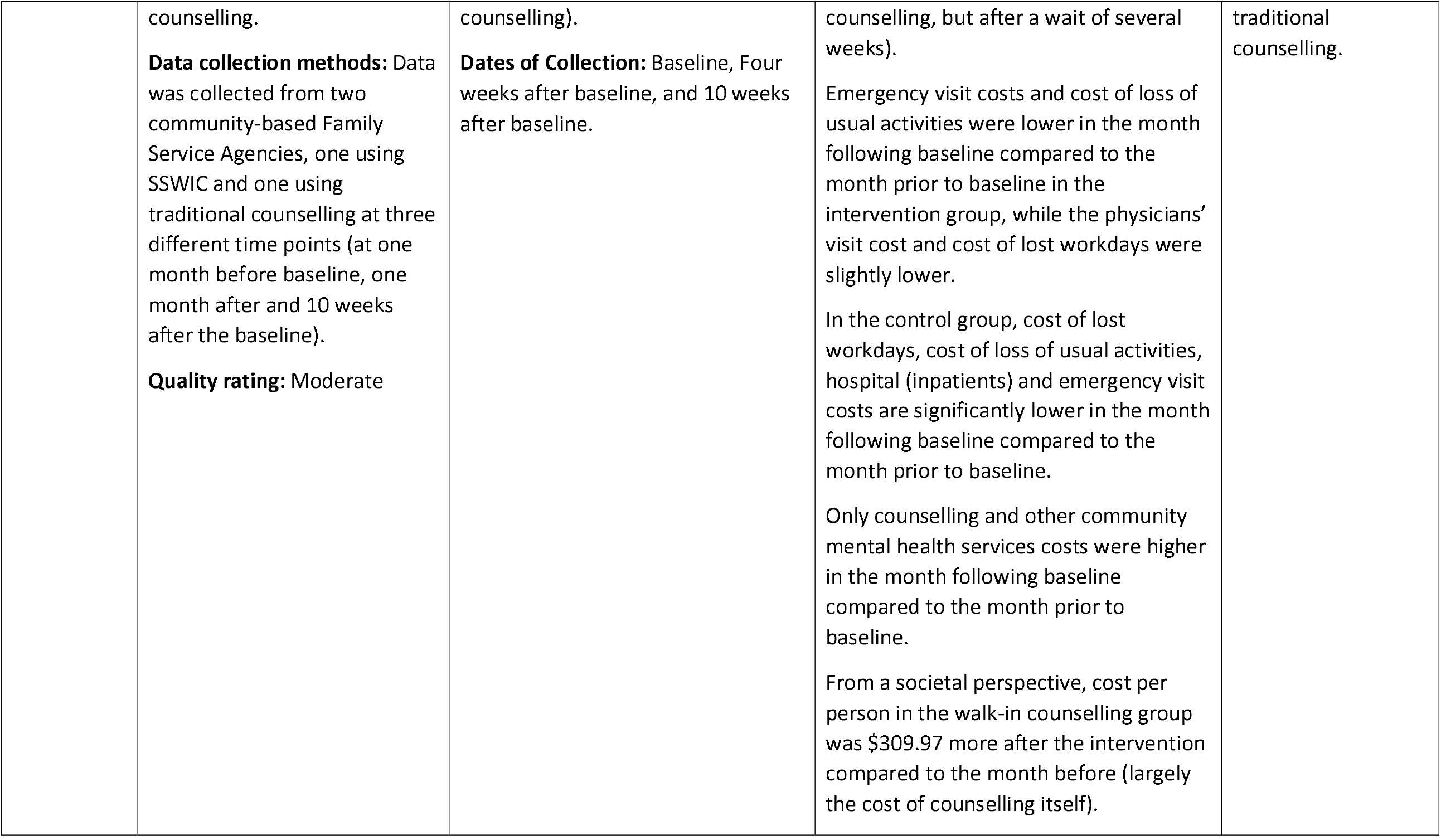
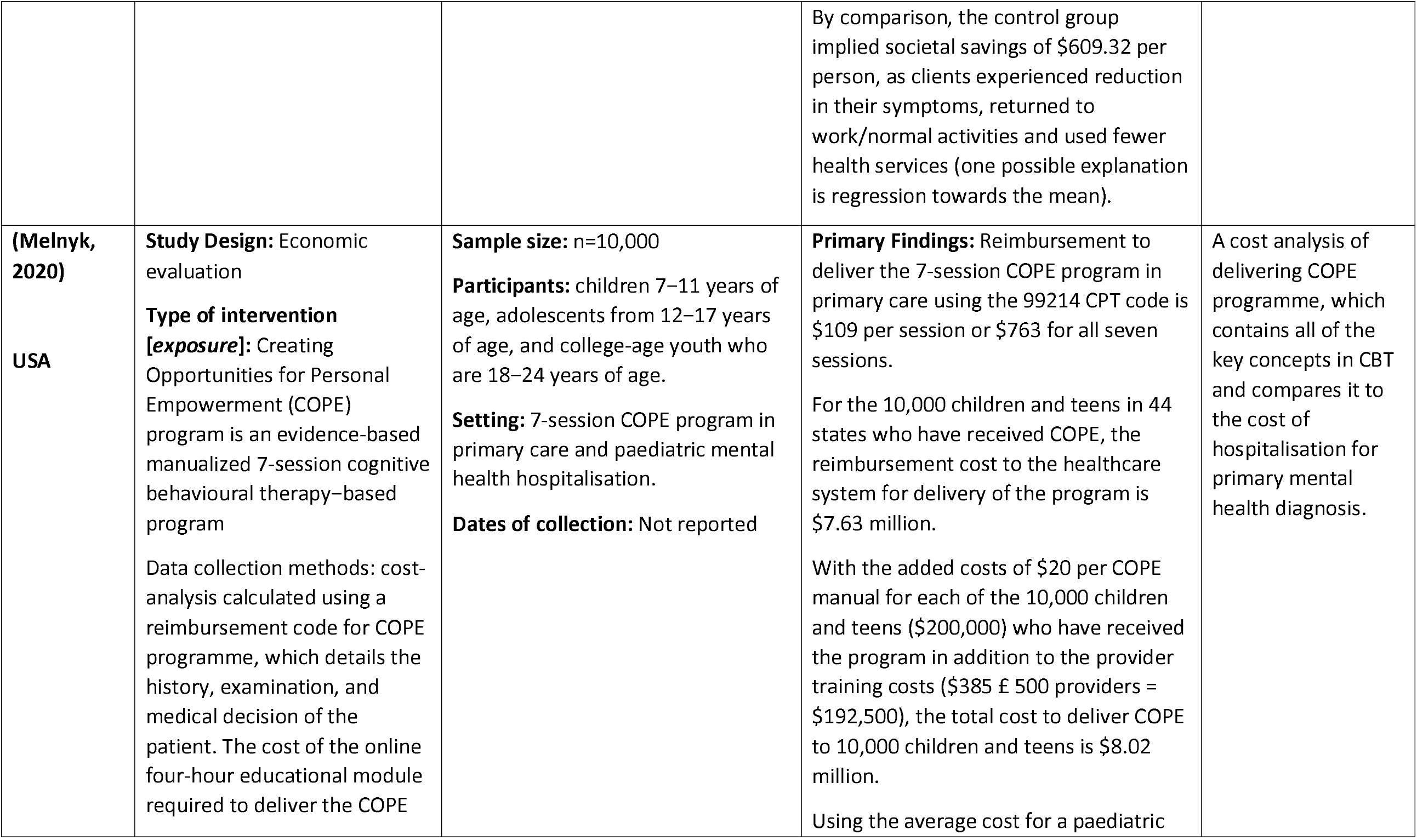
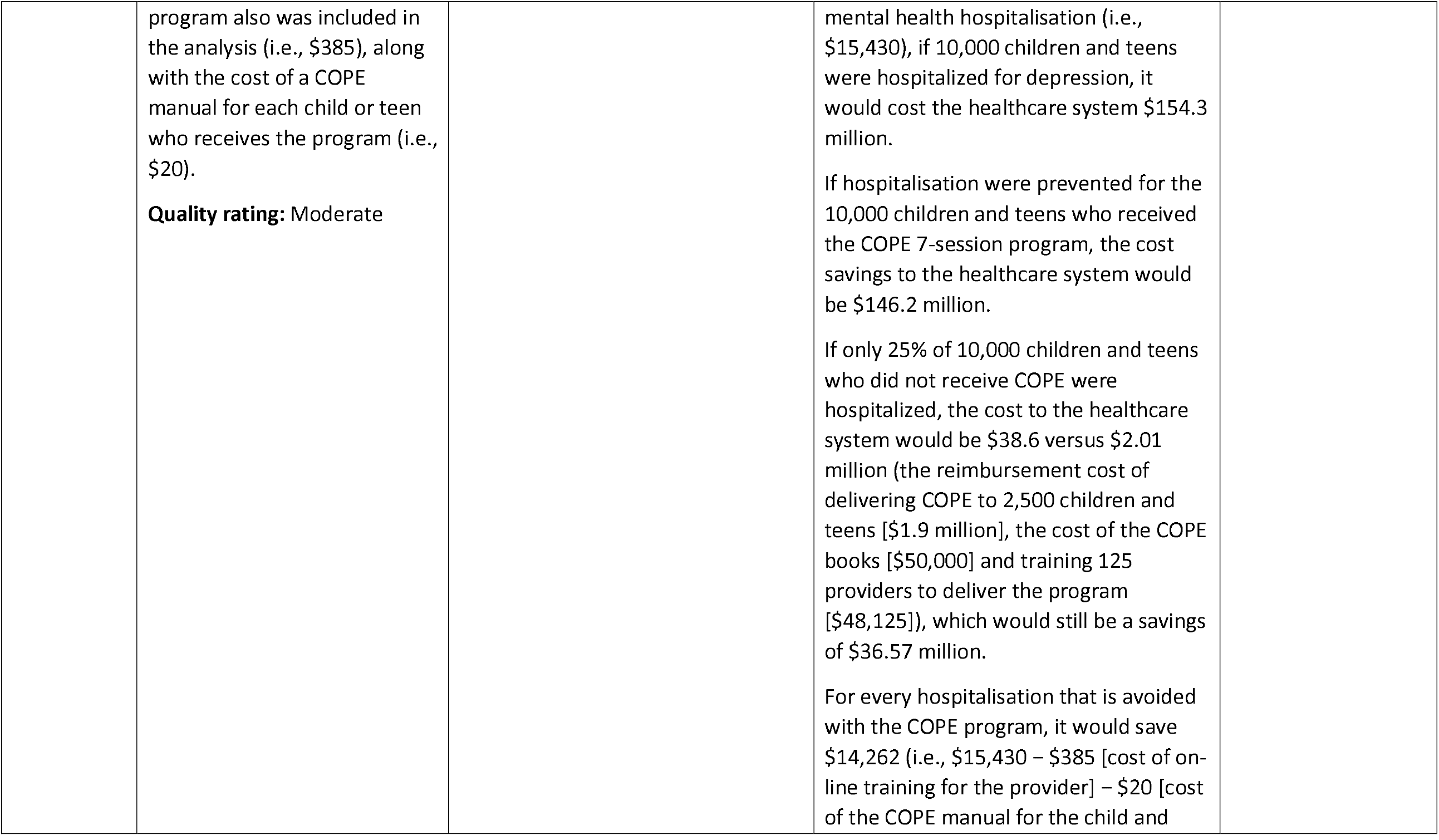
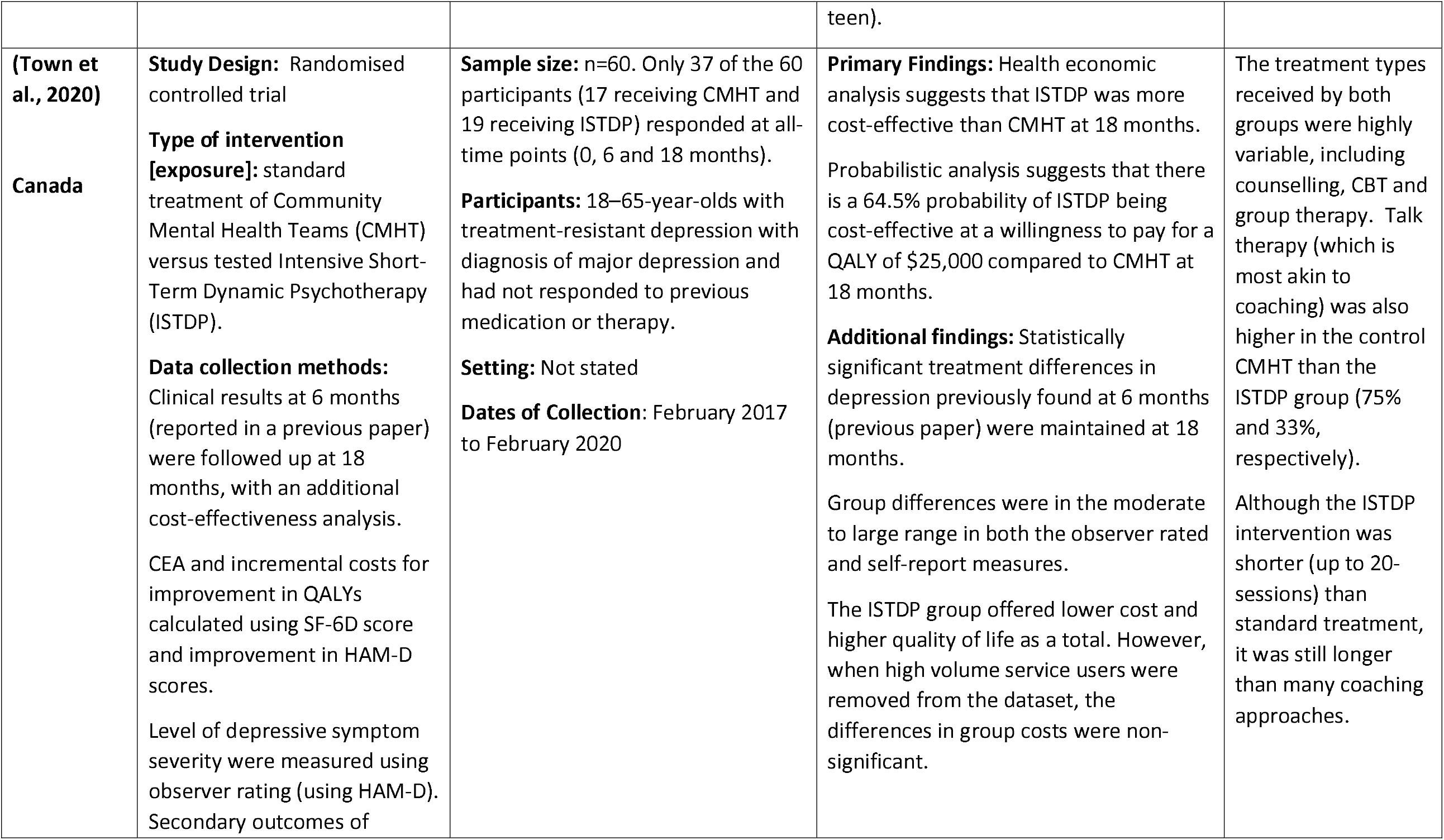
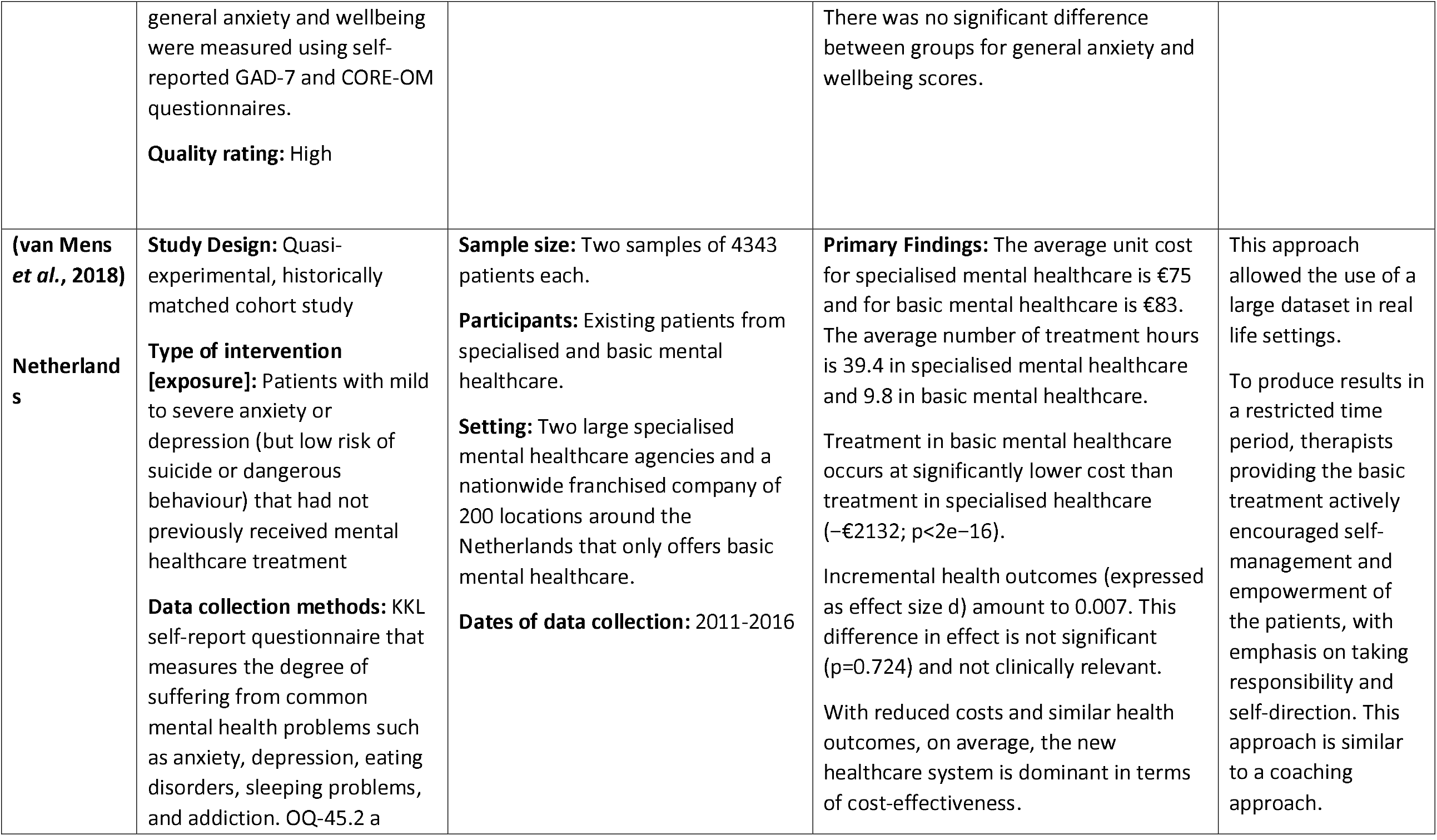
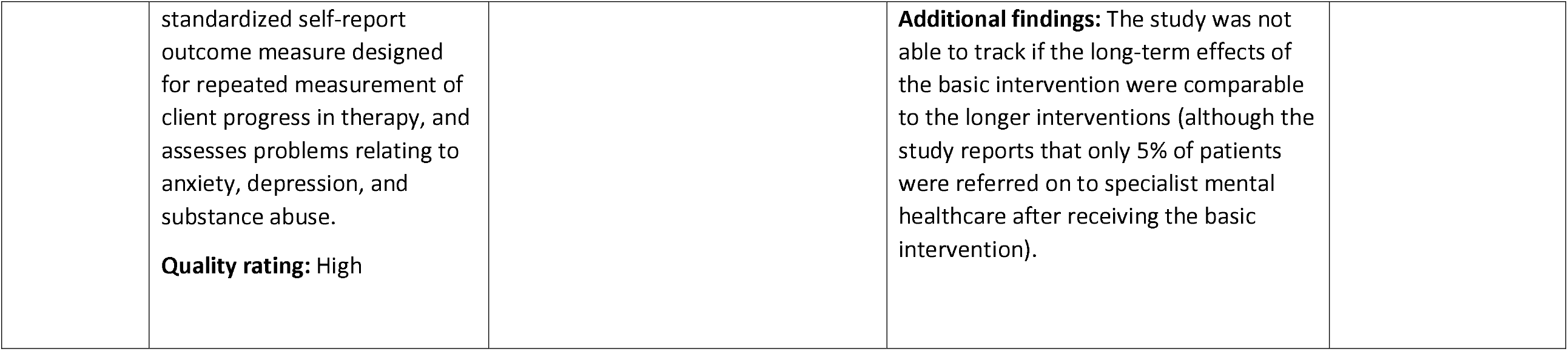

| Citation (Country) | Study Details | Participants & setting | Key findings | Observations/notes |
| --- | --- | --- | --- | --- |
| **(Aas *et al.*, 2019)**<br>**Norway** | **Study Design:** Randomised Clinical Trial<br>**Type of intervention [exposure]:** to evaluate whether trauma-focused cognitive behavioural therapy (TF-CBT) is a cost-effective alternative to therapy as usual (TAU).<br>**Data collection methods:** 15 D Questionnaire to measure HRQoL.<br>The Child Post-Traumatic Stress Scale self-report Questionnaire for Posttraumatic stress symptoms.<br>Mood and Feelings Questionnaire to assess depression<br>Screen for Anxiety Related Emotional Disorders to assess | **Sample size:** n=156<br>**Participants:** Norwegian youth, 10–18 years of age (15.1 years, *SD* = 2.22), presenting with symptoms of posttraumatic stress<br>**Dates of data collection:** 2008 to 2013<br>Five points of participant observation: baseline (T1), after six sessions of treatment (T2), post TF-CBT treatment/after 15 sessions (T3), at a follow-up 1 year after T1 (T4), and at a follow-up 1.5 years after T3 (T5). | **Primary Findings:** TF-CBT is likely to be a cost-effective alternative to standard treatment and should be recommended as the default treatment for youth suffering from PTSD<br>96% of cases of a cost-effectiveness simulation show that TF-CBT is a cost-saving alternative and that the use of other services will decline, such as welfare services, medication, and school nurses. | TF-CBT is a therapy that builds on principles from cognitive, behavioural, interpersonal, and family therapy as well as from trauma-specific interventions. It is a short-term intervention that usually consists of 12 to 15 sessions; however, an extension of up to 30 sessions is recommended for youth with more complex trauma.<br>TAU consists of 53 minutes sessions using therapy strategies which are |
|  | anxiety<br>**Quality rating:** High |  |  | client-centrered, psychodynamic, family, and cognitive intervention. |
| **(De Oliveira et al., 2020)**<br>**USA, Netherlands** | **Study Design:** Systematic Review<br>A before-after case-control or pre-test-post-test component, where measurement of change was present, and include an economic analysis but otherwise was not restricted.<br>**Type of intervention**<br>**[exposure]:** targeting mental health and/or substance use, improve a work-related outcome and be provided in a workplace or be sponsored by the employer.<br>**Quality rating:** High | **Aim:** to review the existing literature and provide a list of the highest yielding and most cost-effective interventions by disorder.<br>**Comparator:** Usual care or no care<br>**Outcomes:** mental health and/or substance use, work-related outcomes, such as productivity, and economic/financial-related outcomes, such as return on investment.<br>**Participants:** all employed adults with a mental health and/or substance use disorder<br>**Dates of data collection:** January 1, 2000 to December 31, 2018 | **Primary Findings:** Moderate evidence from this systematic review indicates that cognitive-behavioural therapy is cost-saving and sometimes cost-effective to address depression.<br>There is robust evidence to suggest that regular and active involvement of occupational health professionals is cost-saving and cost-effective to reduce mental health-related sick leave and encouraging return to work.<br>**Additional Findings:** Dealing with workplace mental health will improve worker wellbeing and productivity and can benefit the employer's profits.<br>56 studies were included out of 11,018 study articles<br>23 studies were of high quality, 25 of fair quality, 7 of poor quality, and 1 of extremely poor quality. | The range of mental health disorders was too vast to synthesise.<br>This systematic review also looked at substance use disorders which was not mentioned in the search criteria. |
| **(Ebert et al., 2018)**<br>**Germany** | **Study Design:** Randomised Clinical Trial<br>To estimate and evaluate the cost-effectiveness and cost-benefit of a guided internet and mobile-supported occupational stress-management intervention (iSMI) for employees from the employer's perspective alongside a randomized controlled trial.<br>The iSMI is an online coaching intervention consisting of seven sessions of problem-solving and emotion-regulation techniques and one booster session. An Online e-coach supports the patients online<br>**Data collection methods:** Perceived Stress Scale (PSS-10).<br>**Quality rating:** High | **Sample size:** n=264<br>**Participants:** employees over the age of 18 with elevated symptoms of perceived stress with a score of 22 or more on the Perceived Stress Scale.<br>**Setting:** a health insurance company in Germany<br>**Dates of data collection:** Online-assessments took place at baseline (T1), post-treatment (seven weeks (T2), and 6-month follow up (T3). | **Primary Findings:** The CBA yielded a net-benefit of €181 [95% confidence interval (CI) -6043–1042] per participant within the first six months following randomization. CEA showed that at a willingness-to-pay ceiling of €0, €1000, €2000 for one additional symptom free employee yielded a 67%, 90%, and 98% probability, respectively, of the intervention being cost-effective compared to the waitlist control group (WLC).<br>**Additional Findings:** The iSMI was cost-effective when compared to WLC and even led to cost savings within the first six months after randomisation. Offering stress-management interventions can present value for money in occupational healthcare. | Both groups had full and unrestricted access to treatment as usual, the WLC only had access to treatment as usual.<br>A limitation of this study is that the contents of treatment as usual were not explicitly outlined. |
| **(Finnes et al., 2017)**<br>**Sweden** | **Study Design:** Randomised Clinical Trial and cost-utility analysis<br>**Type of intervention**<br>**[exposure]:** The evaluation of four interventions: Acceptance and Commitment Therapy (ACT) and workplace dialogue intervention (WDI), both as stand-alone interventions and in combination, compared with treatment as usual (TAU), for employees on sickness absence with mental disorders.<br>**Data collection methods:**<br>Questionnaires and interviews<br>**Quality rating:** High | **Sample size:** n=352 (78.4% female)<br>**Participants:** Employees (working minimum 50% full-term) on sick leave with mental disorders such as anxiety, stress, depression, or adjustment disorder) living in Stockholm County.<br>**Setting:** Swedish places of work<br>**Dates of data collection:** Baseline, post-treatment, 3-months follow up, and 9-months follow up. | **Primary Findings:** ACT is the most cost-effective treatment<br>All four interventions reported HRQoL improvements; however, there were minimal differences between the four interventions in HRQoL and costs.<br>**Additional Findings:** Policy makers should consider ACT as a treatment alternative.<br>TAU and WDI are less economically efficient.<br>The study concluded that WDI should not be added to ACT. | ACT is a sub-category of CBT that focuses on mindfulness, acceptance, and the client's personal values.<br>WDI is a mediation option that seeks to improve communication between the manager and staff. |
| **(Hogewind Johannes et al., 2021)** | **Study Design:** Quasi-experimental research design with cost-effectiveness evaluation. | **Sample size:** 28 divided between the F-ACT intervention (n=13) and TAU (n=15).<br>**Participants:** Adolescents 12-18- | **Primary Findings:** Compared to TAU, F-ACT resulted in a significantly higher quality of life as reported by adolescents.<br>There was no significant difference in |  |
| **Netherlands** | **Type of intervention**<br>**[exposure]:** Youth Flexible Assertive Community Treatment (F-ACT) compared to treatment as usual (TAU).<br>F-ACT youth teams are multidisciplinary including a psychiatrist, a psychologist, a family therapist, case managers, and social workers. F-ACT provides treatment, guidance, and practical assistance with daily living, rehabilitation, and recovery support with to ensure continuity of care, preventing admissions to psychiatric hospitals, and stimulating social functioning and participation.<br>**Data collection methods:**<br>Retrospective cost questionnaire<br>**Quality rating:** High | years of age<br>**Setting:** Community mental healthcare centres<br>**Dates of data collection:** April 2017 to May 2019.<br>Outcomes were assessed at the start and after three months of care.<br>The outcome measures were the percentage of adolescents with a significant improvement in quality of life and Quality Adjusted Life Years (QALYs). | quality of life reported by the parents.<br>The incremental cost-effectiveness ratio (ICER) demonstrated that youth F-ACT is more effective but with higher costs.<br>This study may help to make evidence-based decisions in terms of who benefits the most from youth F-ACT interventions. |  |
| **(Kim et al, 2016)**<br>**South Korea** | **Study Design:** Economic evaluation<br>**Type of intervention [exposure]:** Counselling service<br>**Data collection methods:** Double-bounded dichotomous choice questions.<br>Two survival analysis models were set up with their corresponding variables to investigate the factors affecting the WTP for an increase in insurance premiums to cover counselling services.<br>**Quality rating:** Moderate | **Sample size:** n=561<br>**Participants:** South Korean college students and office workers.<br>**Setting:** South Korean colleges and offices.<br>**Dates of Data Collection:** Not reported | **Primary Findings:** The results indicate that a person in South Korea who makes KRW 2,500,000 (USD 2223.21) per month and pays KRW 66,625 (USD 59.25) per month in insurance premiums is willing to pay a roughly 1% premium to receive additional counselling services.<br>The first regression analysis model for WTP showed that pocket money and counselling experience had significant positive effects while gender had a significant negative effect.<br>The second model included four additional psychological factors and the significant effects of gender and counselling experience that had been found in the first model disappeared while only pocket money showed a significant effect on WTP. | Study utilises the contingent valuation method (CVM) and the willingness to pay (WTP) methods for estimating the economic values of counselling services and identifying the socio-demographic and psychological factors that influence WTP. |
| **(Lamsal et al., 2018)**<br>**Canada** | **Study Design:** Quasi-experimental economic evaluation<br>**Type of intervention [exposure]:** Single session walk in counselling and traditional | **Sample size:** n=524<br>**Participants:** Existing clients aged 16 or over<br>**Setting:** Non-disclosed agency A (Single session walk in counselling and agency B (traditional | **Primary Findings:** The societal and payer costs for SSWIC were higher than for those waiting for traditional counselling, and health outcomes were better. SSWIC is not cost-effective compared to being on the waitlist for traditional counselling (or, for a few patients, having received | Aims to conduct a cost-effectiveness analysis of two models of service delivery, SSWIC compared to being waitlisted for |
| | counselling.<br>**Data collection methods:** Data was collected from two community-based Family Service Agencies, one using SSWIC and one using traditional counselling at three different time points (at one month before baseline, one month after and 10 weeks after the baseline).<br>**Quality rating:** Moderate | counselling).<br>**Dates of Collection:** Baseline, Four weeks after baseline, and 10 weeks after baseline. | counselling, but after a wait of several weeks).<br>Emergency visit costs and cost of loss of usual activities were lower in the month following baseline compared to the month prior to baseline in the intervention group, while the physicians' visit cost and cost of lost workdays were slightly lower.<br>In the control group, cost of lost workdays, cost of loss of usual activities, hospital (inpatients) and emergency visit costs are significantly lower in the month following baseline compared to the month prior to baseline.<br>Only counselling and other community mental health services costs were higher in the month following baseline compared to the month prior to baseline.<br>From a societal perspective, cost per person in the walk-in counselling group was \$309.97 more after the intervention compared to the month before (largely the cost of counselling itself). | traditional counselling. |
| | | | By comparison, the control group implied societal savings of \$609.32 per person, as clients experienced reduction in their symptoms, returned to work/normal activities and used fewer health services (one possible explanation is regression towards the mean). | |
| **(Melnyk, 2020)**<br>**USA** | **Study Design:** Economic evaluation<br>**Type of intervention [exposure]:** Creating Opportunities for Personal Empowerment (COPE) program is an evidence-based manualized 7-session cognitive behavioural therapy-based program<br>Data collection methods: cost-analysis calculated using a reimbursement code for COPE programme, which details the history, examination, and medical decision of the patient. The cost of the online four-hour educational module required to deliver the COPE | **Sample size:** n=10,000<br>**Participants:** children 7–11 years of age, adolescents from 12–17 years of age, and college-age youth who are 18–24 years of age.<br>**Setting:** 7-session COPE program in primary care and paediatric mental health hospitalisation.<br>**Dates of collection:** Not reported | **Primary Findings:** Reimbursement to deliver the 7-session COPE program in primary care using the 99214 CPT code is \$109 per session or \$763 for all seven sessions.<br>For the 10,000 children and teens in 44 states who have received COPE, the reimbursement cost to the healthcare system for delivery of the program is \$7.63 million.<br>With the added costs of \$20 per COPE manual for each of the 10,000 children and teens (\$200,000) who have received the program in addition to the provider training costs (\$385 £ 500 providers = \$192,500), the total cost to deliver COPE to 10,000 children and teens is \$8.02 million.<br>Using the average cost for a paediatric | A cost analysis of delivering COPE programme, which contains all of the key concepts in CBT and compares it to the cost of hospitalisation for primary mental health diagnosis. |
| | program also was included in the analysis (i.e., \$385), along with the cost of a COPE manual for each child or teen who receives the program (i.e., \$20).<br>**Quality rating:** Moderate | | mental health hospitalisation (i.e., \$15,430), if 10,000 children and teens were hospitalized for depression, it would cost the healthcare system \$154.3 million.<br>If hospitalisation were prevented for the 10,000 children and teens who received the COPE 7-session program, the cost savings to the healthcare system would be \$146.2 million.<br>If only 25% of 10,000 children and teens who did not receive COPE were hospitalized, the cost to the healthcare system would be \$38.6 versus \$2.01 million (the reimbursement cost of delivering COPE to 2,500 children and teens [\$1.9 million], the cost of the COPE books [\$50,000] and training 125 providers to deliver the program [\$48,125]), which would still be a savings of \$36.57 million.<br>For every hospitalisation that is avoided with the COPE program, it would save \$14,262 (i.e., \$15,430 – \$385 [cost of on-line training for the provider] – \$20 [cost of the COPE manual for the child and<br>teen). | |
| **(Town et al., 2020)**<br>**Canada** | **Study Design:** Randomised controlled trial<br>**Type of intervention [exposure]:** standard treatment of Community Mental Health Teams (CMHT) versus tested Intensive Short-Term Dynamic Psychotherapy (ISTDP).<br>**Data collection methods:** Clinical results at 6 months (reported in a previous paper) were followed up at 18 months, with an additional cost-effectiveness analysis.<br>CEA and incremental costs for improvement in QALYs calculated using SF-6D score and improvement in HAM-D scores.<br>Level of depressive symptom severity were measured using observer rating (using HAM-D). Secondary outcomes of | **Sample size:** n=60. Only 37 of the 60 participants (17 receiving CMHT and 19 receiving ISTDP) responded at all-time points (0, 6 and 18 months).<br>**Participants:** 18–65-year-olds with treatment-resistant depression with diagnosis of major depression and had not responded to previous medication or therapy.<br>**Setting:** Not stated<br>**Dates of Collection:** February 2017 to February 2020 | **Primary Findings:** Health economic analysis suggests that ISTDP was more cost-effective than CMHT at 18 months.<br>Probabilistic analysis suggests that there is a 64.5% probability of ISTDP being cost-effective at a willingness to pay for a QALY of \$25,000 compared to CMHT at 18 months.<br>**Additional findings:** Statistically significant treatment differences in depression previously found at 6 months (previous paper) were maintained at 18 months.<br>Group differences were in the moderate to large range in both the observer rated and self-report measures.<br>The ISTDP group offered lower cost and higher quality of life as a total. However, when high volume service users were removed from the dataset, the differences in group costs were non-significant. | The treatment types received by both groups were highly variable, including counselling, CBT and group therapy. Talk therapy (which is most akin to coaching) was also higher in the control CMHT than the ISTDP group (75% and 33%, respectively).<br>Although the ISTDP intervention was shorter (up to 20-sessions) than standard treatment, it was still longer than many coaching approaches. |
|  | general anxiety and wellbeing were measured using self-reported GAD-7 and CORE-OM questionnaires.<br>**Quality rating:** High |  | There was no significant difference between groups for general anxiety and wellbeing scores. |  |
| **(van Mens et al., 2018)**<br>**Netherlands** | **Study Design:** Quasi-experimental, historically matched cohort study<br>**Type of intervention [exposure]:** Patients with mild to severe anxiety or depression (but low risk of suicide or dangerous behaviour) that had not previously received mental healthcare treatment<br>**Data collection methods:** KKL self-report questionnaire that measures the degree of suffering from common mental health problems such as anxiety, depression, eating disorders, sleeping problems, and addiction. OQ-45.2 a | **Sample size:** Two samples of 4343 patients each.<br>**Participants:** Existing patients from specialised and basic mental healthcare.<br>**Setting:** Two large specialised mental healthcare agencies and a nationwide franchised company of 200 locations around the Netherlands that only offers basic mental healthcare.<br>**Dates of data collection:** 2011-2016 | **Primary Findings:** The average unit cost for specialised mental healthcare is €75 and for basic mental healthcare is €83. The average number of treatment hours is 39.4 in specialised mental healthcare and 9.8 in basic mental healthcare.<br>Treatment in basic mental healthcare occurs at significantly lower cost than treatment in specialised healthcare (–€2132; $p < 2e-16$).<br>Incremental health outcomes (expressed as effect size d) amount to 0.007. This difference in effect is not significant ($p = 0.724$) and not clinically relevant.<br>With reduced costs and similar health outcomes, on average, the new healthcare system is dominant in terms of cost-effectiveness. | This approach allowed the use of a large dataset in real life settings.<br>To produce results in a restricted time period, therapists providing the basic treatment actively encouraged self-management and empowerment of the patients, with emphasis on taking responsibility and self-direction. This approach is similar to a coaching approach. |
|  | standardized self-report outcome measure designed for repeated measurement of client progress in therapy, and assesses problems relating to anxiety, depression, and substance abuse.<br>**Quality rating:** High |  | **Additional findings:** The study was not able to track if the long-term effects of the basic intervention were comparable to the longer interventions (although the study reports that only 5% of patients were referred on to specialist mental healthcare after receiving the basic intervention). |  |

### Appendix 3: Quality Appraisal Tables

**Table 2:**
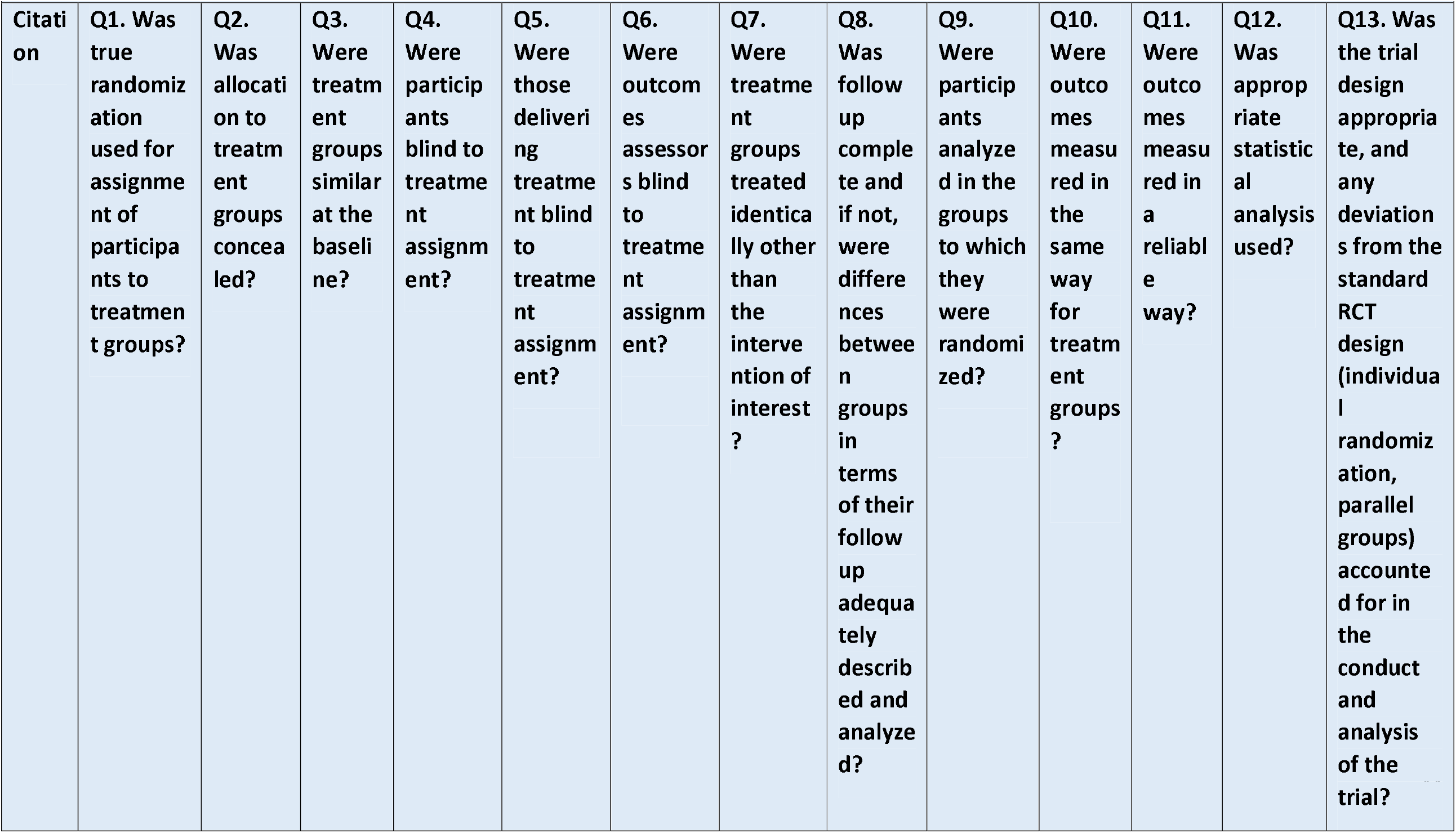
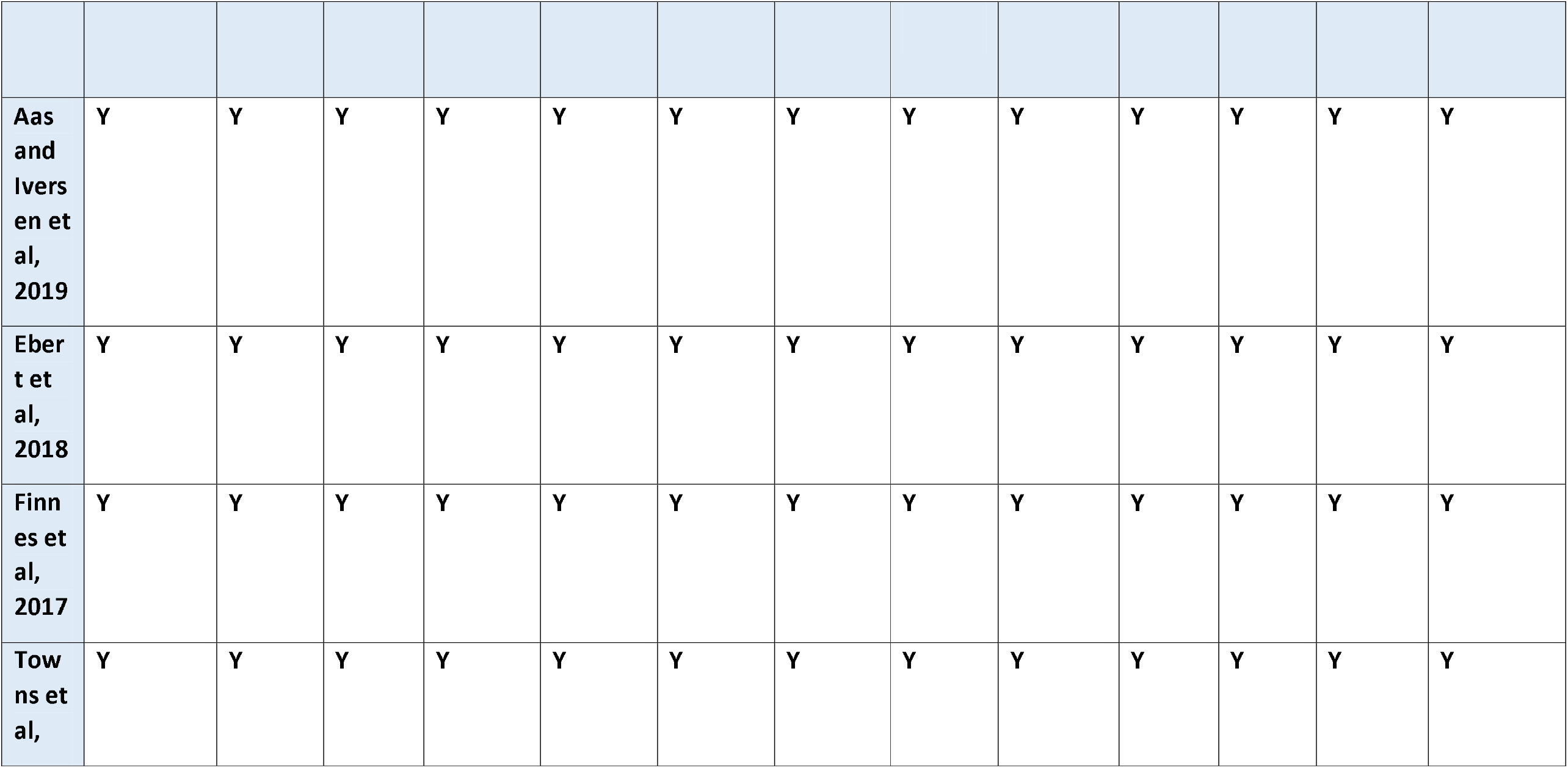
JBI RCT Quality critical appraisal checklist table

**Table 3:**
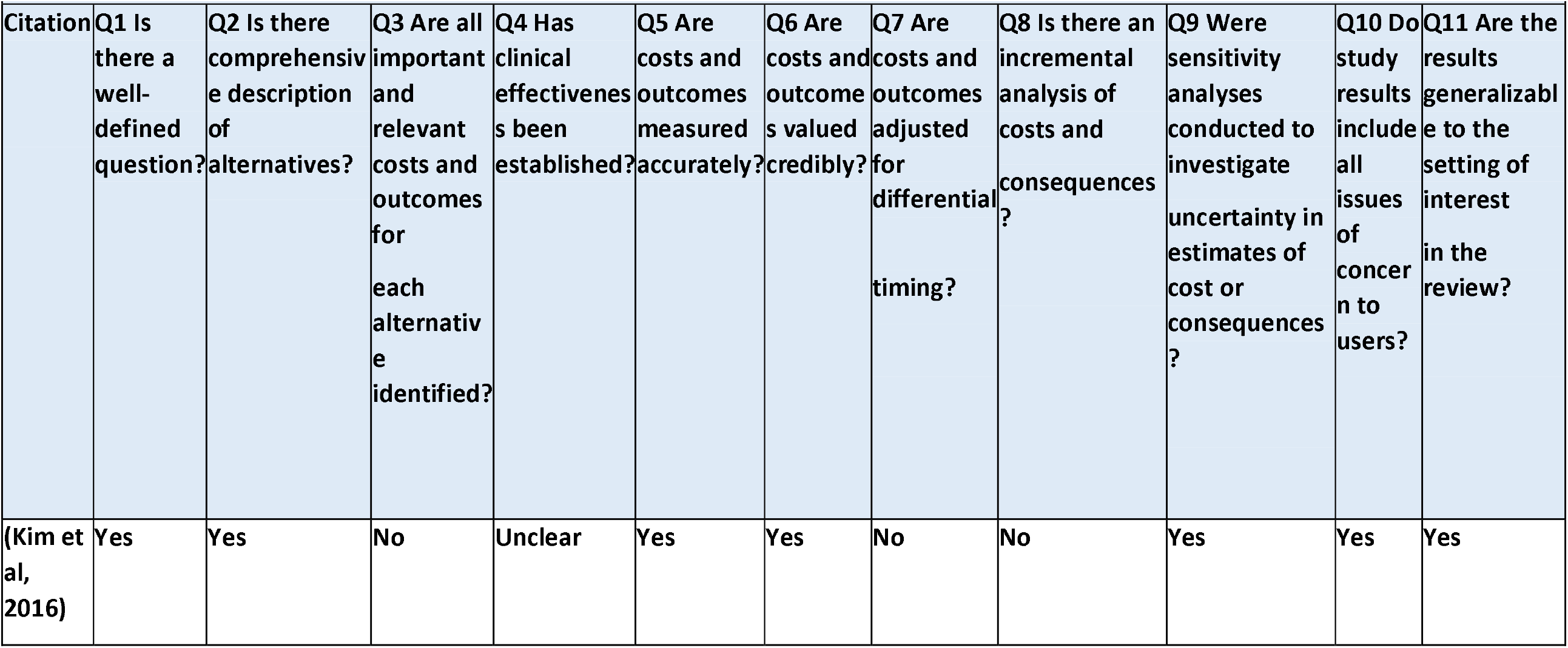
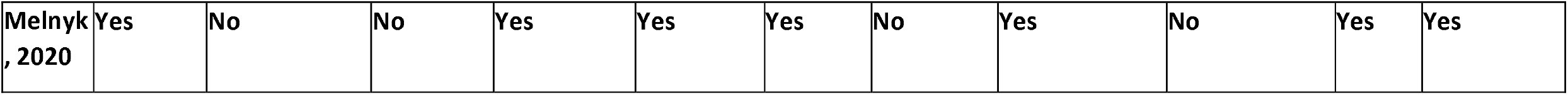
JBI CRITICAL APPRAISAL CHECKLIST FOR ECONOMIC EVALUATIONS

**Table 4:**
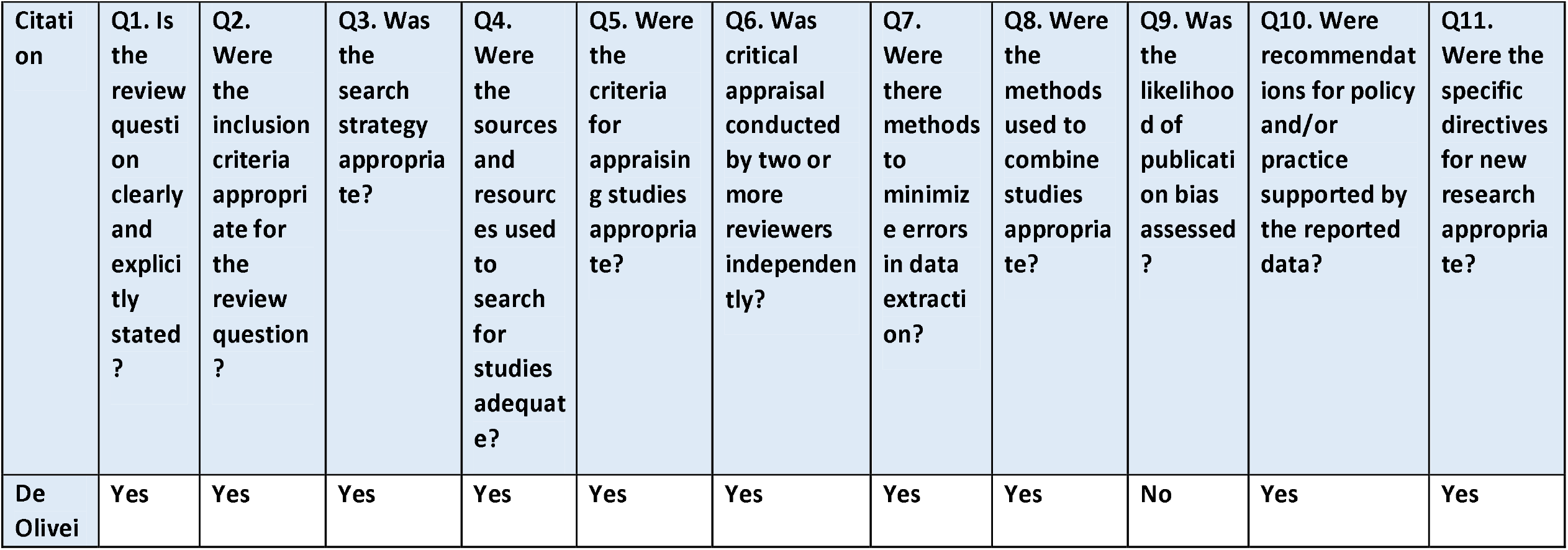
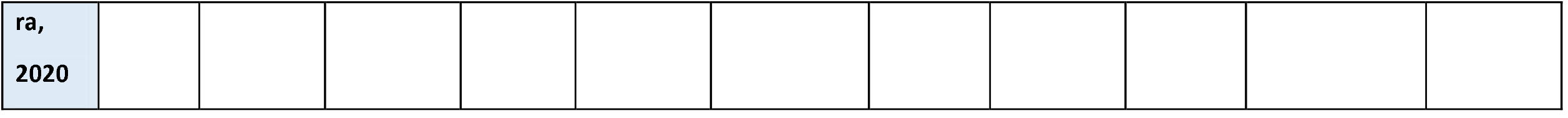
JBI critical appraisal checklist for systematic reviews

**Table 5:**
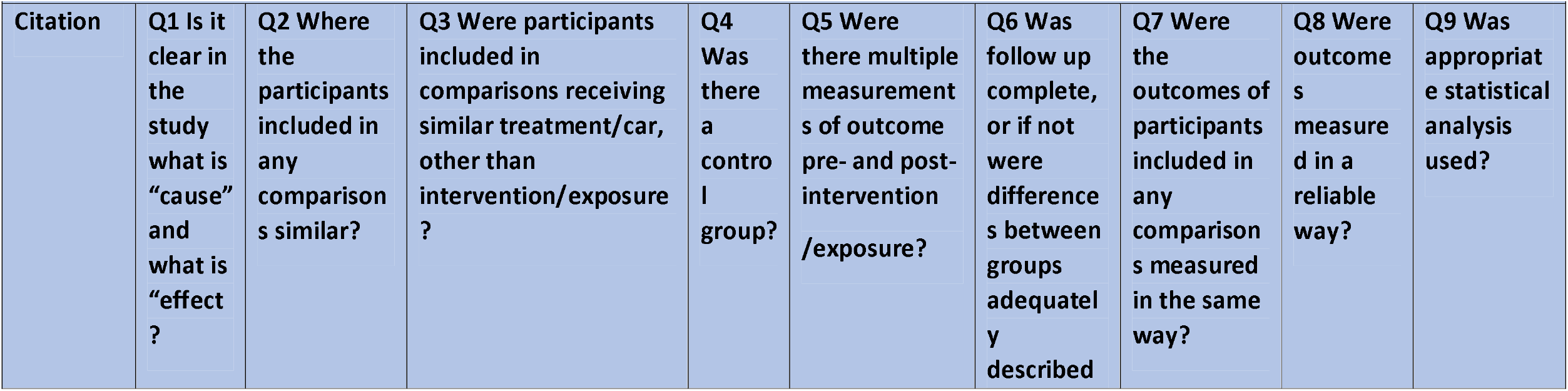
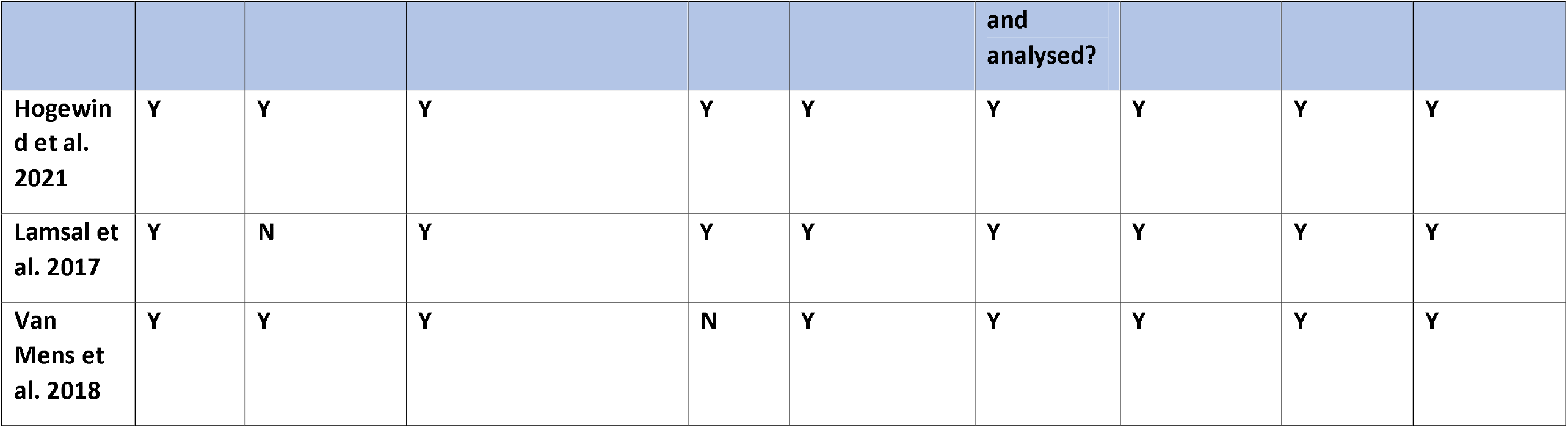
Quasi-Experimental Quality Analysis

| Citation | Q1 Is it clear in the study what is “cause” and what is “effect”? | Q2 Where the participants included in any comparisons similar? | Q3 Were participants included in comparisons receiving similar treatment/control, other than intervention/exposure? | Q4 Was there a control group? | Q5 Were there multiple measurements of outcome pre- and post-intervention/exposure? | Q6 Was follow up complete, or if not were differences between groups adequately described? | Q7 Were the outcomes of participants included in any comparisons measured in the same way? | Q8 Were outcomes measured in a reliable way? | Q9 Was appropriate statistical analysis used? |
| --- | --- | --- | --- | --- | --- | --- | --- | --- | --- |

|  |  |  |  |  |  | and<br>analysed? |  |  |  |
| --- | --- | --- | --- | --- | --- | --- | --- | --- | --- |
| Hogewind et al. 2021 | Y | Y | Y | Y | Y | Y | Y | Y | Y |
| Lamsal et al. 2017 | Y | N | Y | Y | Y | Y | Y | Y | Y |
| Van Mens et al. 2018 | Y | Y | Y | N | Y | Y | Y | Y | Y |

**Abbreviations**

## Abbreviations

ACT: Acceptance and Commitment Therapy
ADHD: Attention deficit hyperactivity disorder
CBA: Cost-Benefit Analysis
CBT: Cognitive Behaviour Therapy
CEA: Cost-effectiveness Analysis
CI: Confidence Interval
CMHT: Community Mental Health Team
CORE-OM: Clinical outcomes routine evaluation - outcome measure
COPE: Creating Opportunities for Personal Empowerment
CVM: Contingent valuation method
GAD-7: Generalised Anxiety Disorder Assessment
HAM-D: Hamilton Depression Rating Scale
HrQoL: Health-Related Quality of Life
iSMI: (guided) internet and mobile-supported occupational stress-management intervention
ISTDP: Intensive Short-Term Dynamic Psychotherapy
JBI: Joanna-Briggs Institute
KRW: South Korean Won
KKL: Korte Klachten Lijst (translates in English to Short Symptom List)
OECD: Organisation for Co-operation and Development
PSS-10: Perceived stress scale
PTSD: Post-traumatic stress disorder
QALY: Quality Adjusted life year
SSWIC: Single Session Walk-in Therapy
TAU: Treatment as Usual
TF-CBT: trauma-focused cognitive behavioural therapy
WDI: Workplace dialogue intervention
WLC: Waitlist Control
WTP: Willingness to pay

